# Reciprocal Feedback Blockade with Trametinib and Imatinib Overcomes the Limitations of Current KRAS-targeted Therapy

**DOI:** 10.64898/2026.07.01.26356985

**Authors:** Yu-Chun Hsiao, Li-Yuan Bai, Yu-Jung Chen, Yi-Syuan Wu, Wei-Jan Wang, Ya-Ling Chuang, Han Chang, Muhammad Zeshan, Heng-Hsiung Wu, Hsiu-Ju Yang, Pei-Chih Lee, Chang-Fang Chiu, Li-Tzong Chen, Hirohito Yamaguchi, Mien-Chie Hung

## Abstract

Although KRAS G12C-specific inhibitors such as sotorasib have been approved by US FDA and currently used in clinic, treating non-G12C mutants and overcoming acquired resistance for these inhibitors remain critical challenges. Here, we introduce a reciprocal feedback blockade therapy combining the MEK inhibitor trametinib and the multi-tyrosine kinase inhibitor imatinib to overcome these limitations. Our study reveals their compensatory roles: trametinib suppresses MEK activity yet promotes tyrosine kinase signaling and angiogenesis, while imatinib, a pan-tyrosine kinase inhibitor unleashes the MEK/ERK pathway via phosphatase suppression. Combining these agents blocks the reciprocal survival signals, inducing robust cell death across diverse *KRAS*-mutant models. Mechanistically, this combination reprograms cellular metabolism, leading to autophagy-dependent lipid peroxidation accumulation and ferroptosis. This strategy was effective in sotorasib-resistant lung cancer cells and various mouse models, including pancreatic cancer patient-derived xenograft. Furthermore, a pilot clinical trial for *KRAS*-mutant pancreatic cancer yielded encouraging responses. Consequently, the trametinib-imatinib combination represents a promising, broad-spectrum therapeutic strategy to overcome the constraints of current KRAS-targeted therapies.

## Introduction

Kirsten rat sarcoma 2 viral oncogene homolog (*KRAS*) is one of the most frequently mutated oncogenes in solid tumors, including pancreatic ductal adenocarcinoma (PDAC), colorectal cancer (CRC), and non-small cell lung cancer (NSCLC)^1,2^. The majorly of *KRAS* mutations occur at codon G12 (G12D, G12V, G12C, G12R, etc.) and act as potent drivers of tumor initiation, progression, and therapeutic resistance, highlighting KRAS as a key focus in cancer research and targeted therapy development^3–6^.

Recently, sotorasib and adagrasib, KRAS G12C-specific inhibitors, have received U.S. Food and Drug Administration (FDA) approval for treatment of patients with *KRAS* G12C-mutated NSCLC ^7,8^. However, the objective response rates of these drugs are approximately 30∼40 % and the median progression-free survival (PFS) is around 6 months. Furthermore, acquired drug resistance emerges in many cases, suggesting that theses therapies remain an early approach rather than a definitive cure ^9–12^. Moreover, these drugs are ineffective against tumors harboring non-G12C *KRAS* variants, leaving a substantial unmet medical need. Although several compounds targeting other *KRAS* mutations have been developed and are currently under clinical investigation, none have yet achieved U.S. FDA approval^13–15^.

The MAP kinase pathway (MEK-ERK) is a major downstream RAS signaling pathway, and the MAPK pathway has emerged as a central therapeutic target in clinical oncology. Trametinib, a selective MEK1/2 inhibitor, has been approved in combination with dabrafenib for *BRAF* V600E-mutated NSCLC and melanoma and is currently under evaluation in phase II clinical trials for PDAC ^16–18^. However, its efficacy as monotherapy for *KRAS*-mutated cancers remains limited^19^. Furthermore, combination therapies with targeted therapy^20–22^, chemotherapy^18,23^ or immunotherapy^24–26^ have also shown limited efficacy and often significant adverse events. These limitations highlight the need for deeper understanding of compensatory pathways and resistance mechanisms to MAPK inhibition in *KRAS*-mutated cancers, and for development of new therapeutic strategies based on these mechanisms.

This study stemmed from an unexpected observation that a MEK inhibitor activates multiple receptor tyrosine kinases (RTKs) and angiogenesis in *KRAS*-mutated but not *KRAS* wild-type (WT) cancer cells and tumors, respectively. We then screened tyrosine kinase inhibitors with anti-angiogenic activity for combining with a MEK inhibitor. We found that combining the multi-tyrosine kinase inhibitor imatinib and MEK inhibitor trametinib showed a potent inhibitory effect on various *KRAS*-mutated cancer cells. Furthermore, we showed that the combination therapy of trametinib and imatinib contributed to improved antitumor activity by promoting ferritinophagy and subsequent ferroptosis through reprogramming the metabolism of tumor cells. This combination therapy showed potent efficacy in clinically relevant mouse models such as *KRAS*-mutated orthotopic xenograft models and patient-derived xenograft (PDX) models. Furthermore, to evaluate the clinical potential of the combination therapy in patients with *KRAS*-mutated solid tumors, a pilot investigator-initiated trial (pilot-IIT) (NCT06962254) was conducted, in which the combination therapy of trametinib and imatinib demonstrated high tolerability and durable disease stabilization in patients with advanced PDAC with non-*KRAS* G12C mutations. Taken together, these findings suggest that simultaneously targeting multi-tyrosine kinases and the MEK/ERK signaling pathway could be a highly promising therapeutic strategy for a wide range of *KRAS*-mutated cancers.

## Result

### Adaptive activation of multiple receptor tyrosine kinases and angiogenic programs limits the efficacy of trametinib in KRAS-mutated cancer cells

Constitutive activation of the MEK/ERK is a hallmark of *KRAS*-mutated malignancies, including PDAC and NSCLC. Therefore, MEK inhibitors have been extensively explored in the clinical trials as targeted therapies for *KRAS* mutant cancers, but their efficacy has been limited. One mechanism of resistance to MEK inhibition in *BRAF*-mutated cancers has been shown to be activation of signaling via upregulation of RTKs^27–29^. Therefore, to evaluate the upregulation of tyrosine kinase activity after trametinib treatment in *KRAS*-mutated cell lines, we performed RNA sequencing followed by gene set enrichment analysis (GSEA) in trametinib-treated and untreated *KRAS*-mutated A549 cells. Indeed, trametinib treatment resulted in significant enrichment of gene signatures associated with RTK signaling in *KRAS* mutant cells (Fig. 1a). Moreover, differentially expressed gene (DEG) analysis revealed coordinated transcriptional remodeling of RTK pathways, including increased expression of IGFR-signal related molecules, *FGFR1*, *EGFR*, *PDGFRA/B*, and *ERBB2*, alongside reduced expression of *FGFR2*, *FGFR3*, *FGFR4*, *ERBB3*, *AXL* (Extended Data Fig. 1a). To assess whether these transcriptional changes lead to functional RTK activation, we performed a human RTK antibody array in trametinib-treated and untreated CFPAC cells, and found that trametinib treatment increased phosphorylation of multiple RTKs, including EGFR, ErbB2, FGFR1, IGF-1R, HGFR (c-MET), and PDGFRα (Extended Data Fig. 1b).

**Fig. 1.**
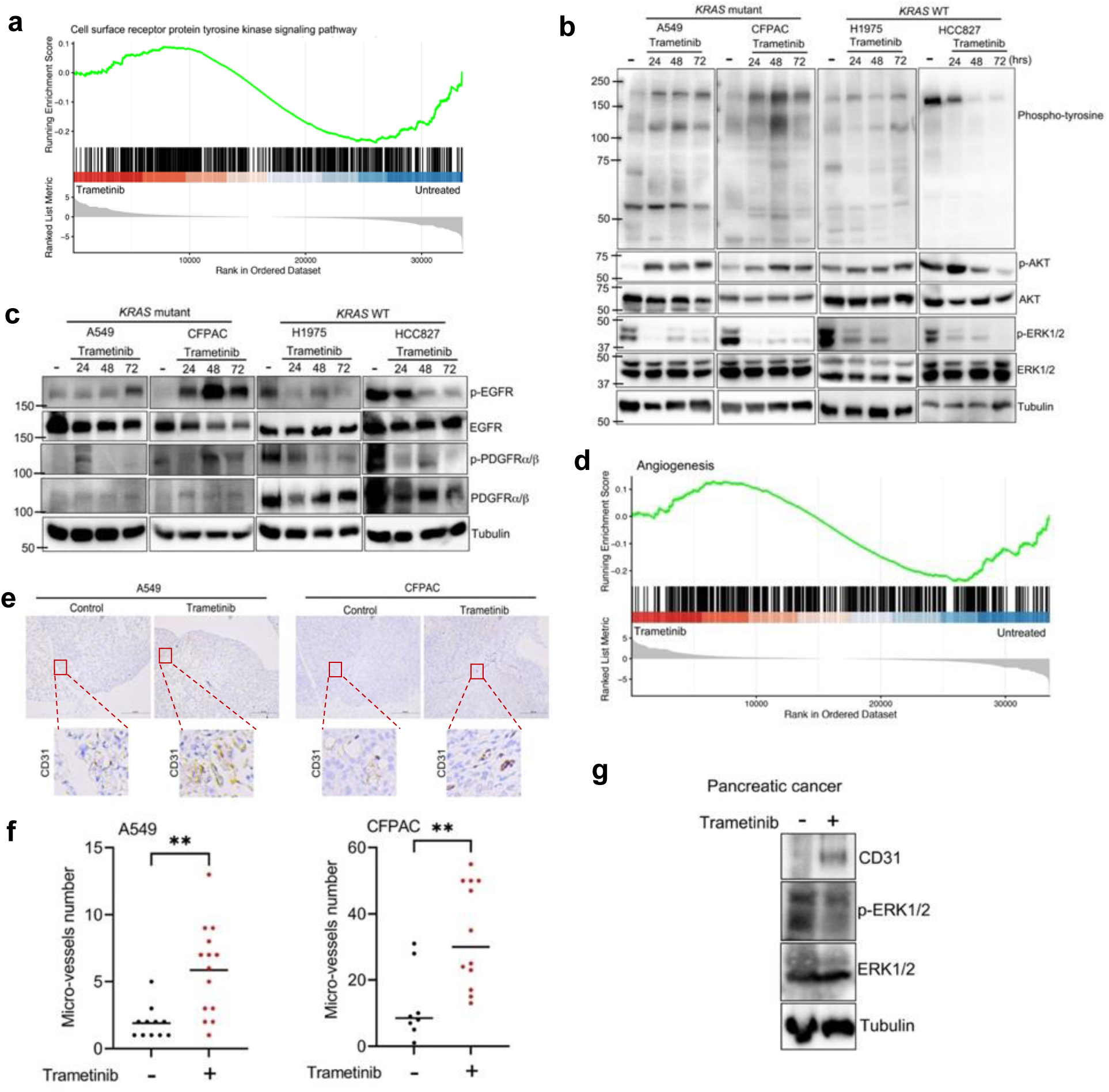
Adaptive activation of receptor tyrosine kinases and an angiogenic program by trametinib in *KRAS* mutant cancer cells. **a**, Gene set enrichment analysis (GSEA) of receptor tyrosine kinase (RTK)-related gene sets using RNA-seq data from A549 cells treated with trametinib (30 nM) or vehicle for 24 hours. **b,** *KRAS* mutant A549 and CFPAC cells and KRAS WT H1975 and HCC827 were treated with trametinib (30 nM) for the indicated times, and their cell lysates were analyzed by western blot to detect phosphorylated tyrosine kinases, p-AKT (S473), AKT, p-ERK1/2 (Y204/T202) and ERK1/2. **c,** Western blot analysis of Y1068-phosphorylated EGFR (p-EGFR), EGFR, Y849-phosphorylated PDGFR⍺/Y857-phosphorylated PDGFRβ (p-PDGFR⍺/β), PDGFR⍺/β, in *KRAS* mutant A549 and CFPAC cells and *KRAS* WT H1975 and HCC827 cells treated with trametinib (30 nM) for indicated times. **d,** GSEA of angiogenesis-related gene sets using RNA-seq data from A549 cells treated with trametinib (30 nM) or vehicle for 24 hours. **e,** Representative immunohistochemical (IHC) images (4×) of the angiogenesis marker CD31 in tumors from *KRAS* mutant A549 lung cancer xenografts and *KRAS* mutant CFPAC pancreatic cancer xenografts treated with trametinib for 30 days. **f,** Quantification of CD31 IHC staining shown in (**e**). Data represent mean ± SD from ten mice. **p < 0.01. **g,** Western blot analysis of CD31, p-ERK1/2 (Y204/T202), and ERK1/2 in tumor lysates from *KRAS* mutant pancreatic cancer xenografts treated with trametinib (0.15mg/kg).

To further verify these results, we treated *KRAS* mutant cell lines A549 and CFPAC, and *KRAS* WT cell lines H1975 and HCC827 with trametinib for up to 3 days and subjected them to western blot analysis. Trametinib effectively suppressed MEK/ERK signaling in both *KRAS* mutant and *KRAS* WT cells as expected, but un-expectedly selectively induced an increase in phosphotyrosine levels accompanied by AKT activation only in *KRAS* mutant cells (Fig. 1b). Consistently, time-course analyses by western blotting validated a progressive of RTK activation following trametinib treatment, with prominent phosphorylation of EGFR at Y1068 (p-EGFR) and PDGFRα/β at Y849/Y857 (p-PDGFR α/β) observed selectively in *KRAS* mutant A549 and CFPAC cells (Fig. 1c). In contrast, trametinib suppressed p-EGFR and p-PDGFRα/β in *KRAS* WT H1975 and HCC827 cells.

In addition, it has been known that the PI3K-AKT pathway is critical for RTK signaling and resistance to KRAS pathway inhibition^30,31^. Consistent with this, trametinib treatment markedly induced AKT S473 phosphorylation (p-AKT) in *KRAS* mutant A549 and CFPAC cells (Fig. 1b). In contrast, AKT activation was not induced by trametinib in *KRAS* WT H1975, but was also effectively suppressed in *KRAS* WT HCC827 cells (Fig. 1b). Furthermore, analysis of RNA sequencing data revealed upregulation of AKT-associated genes including *EGFR*, *PDGFRA* and *PDGFRB* following trametinib treatment selectively in *KRAS* mutant cells (Extended Data Fig. 1a and Extended Data Fig. 1c). These findings suggest that RTK and AKT reactivation may contribute to intrinsic resistance to MEK inhibition in *KRAS* mutant cancers.

Since PDGFR and EGFR are known to strongly induce angiogenesis, and both were activated in *KRAS-*mutated cancer cells treated with trametinib, we asked if trametinib may promote angiogenesis in *KRAS*-mutated tumors. Indeed, the result of the GSEA suggested that MEK inhibition promotes angiogenic programs in *KRAS* mutant tumors (Fig. 1d). Therefore, we examined micro-vessel density using CD31 immunohistochemistry (IHC) staining in *KRAS*-mutated lung and pancreatic cancer xenograft mouse models treated/untreated with trametinib. CD31 staining was significantly increased in both xenograft tumors treated with trametinib compared to tumors in the untreated group (Fig. 1e, f). Consistent with these findings, western blot analysis of tumor lysates verified increased CD31 expression despite effective suppression of MEK/ERK signaling following trametinib treatment (Fig. 1g). Collectively, these results demonstrate that MEK inhibition in *KRAS* mutant cancers induces compensatory activation of multiple RTKs, AKT signaling, and angiogenic programs, providing a mechanistic explanation for the limited efficacy of trametinib monotherapy.

### Combination therapy with imatinib enhances the sensitivity of KRAS-mutated cells to MEK pathway inhibition

The results shown above suggest that drugs possessing both tyrosine kinase inhibition and anti-angiogenic effects are promising candidates for combination therapy with MEK inhibitors for *KRAS* mutated cancers. To this end, we first selected several candidate compounds from the library of 700 FDA-approved compounds. These drugs were prioritized based on their dual functions, including tyrosine kinase inhibition and anti-angiogenic activity.

We next examined the effects of selected candidate drugs on AKT and ERK signaling, main downstream signaling pathways of KRAS, in *KRAS* mutant A549 cells and *KRAS* WT HCC827 cells. Among the screened compounds, imatinib most effectively suppressed p-AKT in A549 cells, but not in HCC827 cells (Fig. 2a). Imatinib is an inhibitor of multiple tyrosine kinases such as ABL, c-KIT, and PDGFRα/β and was the first small-molecule tyrosine kinase inhibitor approved by the U.S FDA in 2002 for treatment chronic myeloid leukemia (CML). Imatinib is also shown to inhibit VEGF-independent angiogenesis^32^. Interestingly, imatinib effectively suppressed AKT in A549 cells, but it did not induce apoptosis as assessed by PARP cleavage. This would be attributed to the paradoxical enhancement of p-ERK1/2 (the antibody recognizes both Y204/T202 phosphorylation) after imatinib treatment (Extended Data Fig. 2a).

**Fig. 2:**
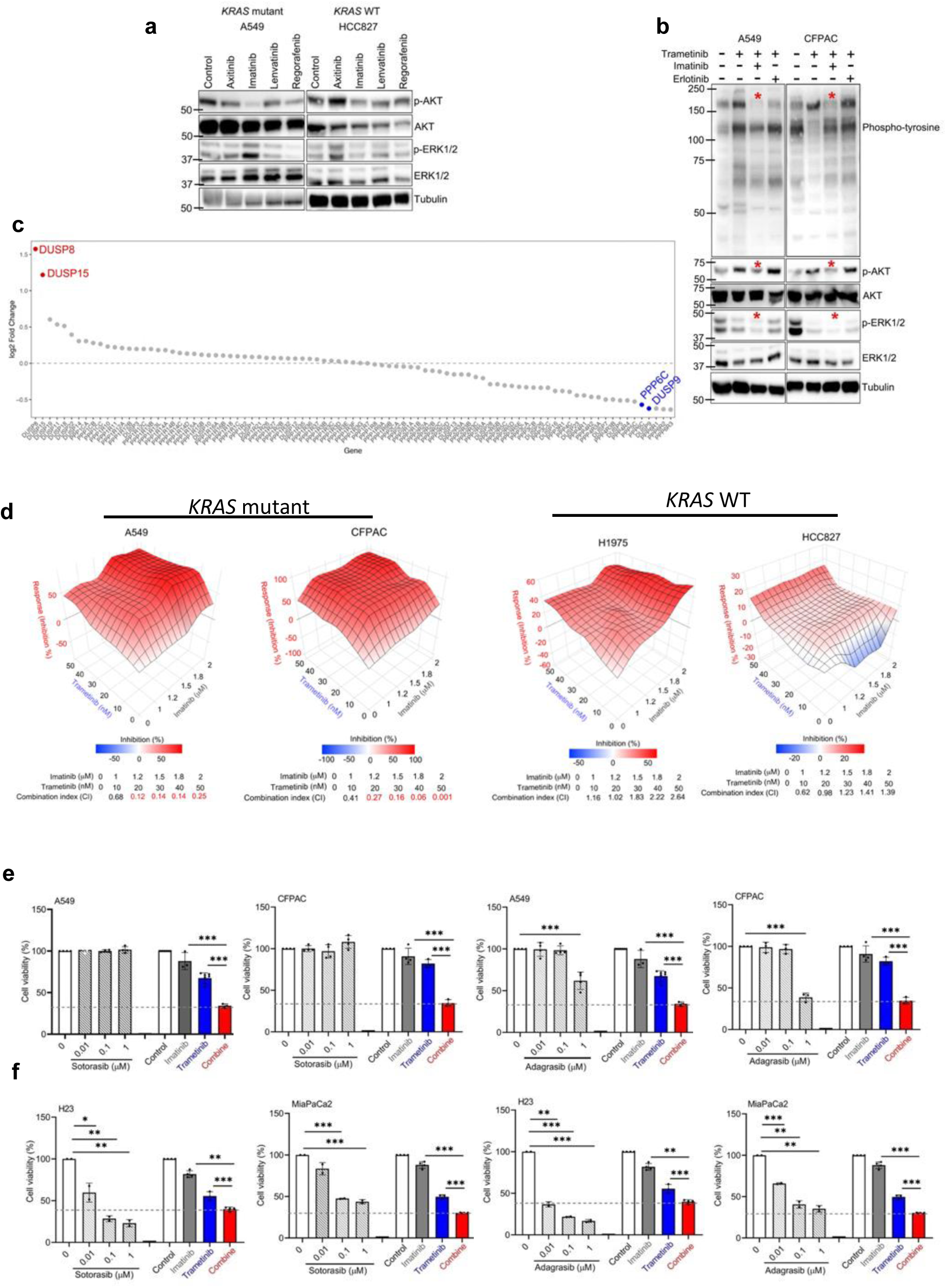
Identification of imatinib as a synergistic partner with trametinib in *KRAS* mutant cells. **a**, Western blot analysis of p-AKT (S473), AKT, p-ERK1/2 (Y204/T202), and ERK1/2 in A549 and HCC827 cells treated for 24h with 1 µM axitinib, 1µM imatinib, 1 µM lenvatinib or 1 µM regorafenib. **b,** Western blot analysis of phosphorylated tyrosine kinases, p-AKT (S473), AKT, p-ERK1/2 (Y204/T202), and ERK1/2 in A549 and CFPAC following treatment with trametinib alone or in combination with imatinib (1 μM) or erlotinib (1 μM). **c,** Dot plot showing log₂ fold changes in phosphatase gene expression between the combination-treated and untreated A549 cells. **d,** Colony formation assays demonstrating synergistic anti-cancer effects of trametinib and imatinib in *KRAS* mutant A549 and CFPAC cells, as well as *KRAS*-WT H1975 and HCC827 cells treated with indicated concentrations for 6 days. **e,** Colony formation assay of A549 and CFPAC cells treated for 6 days with sotorasib, adagrasib, imatinib (1 µM), trametinib (30 nM), or imatinib-trametinib combination at the indicated concentrations. Data represent mean ± SD from at least three independent experiments. ***p < 0.001. **f,** Colony formation assays of H23 and MiaPaCa2 cells treated as described in (e). Data represent mean ± SD from at least three independent experiments. *p < 0.05, **p < 0.01, ***p < 0.001.

We extended these observations to additional *KRAS*-mutant models, including NSCLC cell lines H23 (*KRAS* ^G12C^), H441 (*KRAS* ^G12V^), A549 (*KRAS* ^G12S^) as well as PDAC cell lines MiaPaCa-2 (*KRAS* ^G12C^), CFPAC (*KRAS* ^G12V^), and SU.86.86 (*KRAS* ^G12D^). Imatinib treatment suppressed p-AKT (S473) in A549 and CFPAC cells, while simultaneously enhancing p-MEK1/2 (the antibody recognizes both S217/221 phosphorylation) and p-ERK1/2 across multiple *KRAS* mutant lines (Extended Data Fig. 2b, c). In contrast, imatinib did not activate MEK1/2, or ERK1/2 in *KRAS* WT H1975 and HCC827 cells (Extended Data Fig. 2b, c). BRaf and CRaf are the Raf family kinases, which was the critical components of the RAF-MEK-ERK signaling pathway. Among screened these components, imatinib treatment did not affect BRaf S445 phosphorylation (p-BRaf) and CRaf S338 phosphorylation (p-CRaf) in *KRAS* mutant and WT cells (Extended Data Fig. 2b). These findings are consistent with prior reports demonstrating imatinib-induced ERK activation is a compensatory mechanism limiting therapeutic efficacy in *KRAS* mutant PDAC cells^33^. Moreover, because YAP/TAZ activation has been implicated in resistance to KRAS pathway inhibition, we also examined whether imatinib modulated YAP/TAZ activation ^34,35^. Imatinib treatment did not alter YAP/TAZ activation in *KRAS* mutant NSCLC or PDAC models (Extended Data Fig. 2d), suggesting that alternative feedback mechanism underlies ERK reactivation.

To identify molecular mediators of imatinib-induced MAPK activation in *KRAS*-mutated cancer cells, we performed RNA sequencing in *KRAS* mutant cells treated or untreated with imatinib. Transcriptome analysis revealed downregulation of multiple dual-specificity phosphatases (DUSPs) and phosphoprotein phosphatases (PPPs) following imatinib treatment (Extended Data Fig. 2e). qPCR validation confirmed consistent downregulation of *DUSP18*, *PPP1R3C*, *DUSP8*, *PPP3CA*, *PPP1R21*, *DUSP15*, and *PPP1R12A* in *KRAS* mutant cells, whereas these transcripts were upregulated in *KRAS* WT cells (Extended Data Fig. 2f). Western blotting validated that DUSP8 and DUSP15 proteins were consistently downregulated in *KRAS* mutant cells (Extended Data Fig. 2g). Overexpression of either DUSP8 or DUSP15 but not PPP1R21 in 293T cells suppresses p-MEK1/2 and p-ERK1/2 (Extended Data Fig. 2h–2j), and similar effect was observed in A549 and CFPAC cells (Extended Data Fig. 2k-2m). These findings indicate that imatinib-induced downregulation of DUSP8 and DUSP15 drives feedback activation of the MEK/ERK pathway, thereby limiting imatinib sensitivity in *KRAS* mutant cancer cells.

Considering that imatinib most potently suppresses AKT activity among the drugs tested, and that imatinib-induced MEK/ERK activation can be suppressed by trametinib, combination therapy with imatinib and trametinib appears to be a promising strategy for *KRAS*-mutated cancers. Therefore, we first investigated the effects of the combination of imatinib and trametinib on tyrosine kinase levels and AKT and ERK activation (Fig. 2b). In *BRAF*-mutated cancers, EGFR activation is also involved in resistance to MEK inhibition, and combination therapy with EGFR inhibitors and MEK inhibitors has been investigated in several preclinical studies ^36,37^. Thus, as a control, we used the EGFR inhibitor erlotinib and trametinib in combination. Interestingly, only the combination of trametinib with imatinib, but not the trametinib with erlotinib, effectively suppressed phosphotyrosine signaling, AKT and ERK activation (Fig. 2b). Further analysis of RNA sequencing results revealed that suppression of gene expression including MAPK family (*MAP2K1*, *MAP2K2*, *MAP2K3*, *MAP2K4*, *MAP2K5*, *MAP2K7*, and *MAP2K6*), AKT signaling (*AKT1*, and *AKT2*), and tyrosine kinase family (*EGFR*, *FGFR2*, *FGFR3*, and *FGFR4*), following combination treatment (Extended Data Fig. 2n). Interestingly, it was shown that *DUSP8* and *DUSP15* transcripts were selectively upregulated by combination therapy in *KRAS* mutant cells, suggesting that the restoration of these phosphatase suppression may contributes to of imatinib-induced MEK/ERK activation in the combined treatment (Fig. 2c).

Next, to evaluate the synergistic effects of the combination of imatinib and trametinib, we evaluated cell viability after 6 days of single or combination treatment in *KRAS* mutant cells including A549, H441, CFPAC, and SU.86.86 and *KRAS* WT cell lines including H1975 and HCC827, and the combination index (CI) was determined. The results revealed strong synergistic effects (CI ≤ 0.3) in all the *KRAS* mutant cells, but not in the *KRAS* WT cells (Fig. 2d, Extended Data Fig. 3a and Extended Data Fig. 3b).

*KRAS* G12C inhibitors, including sotorasib and adagrasib, have been approved for *KRAS* G12C mutant NSCLC. Therefore, we compared the efficacy of trametinib plus imatinib with the *KRAS* G12C inhibitors in *KRAS* mutant models using a colony formation assay. As expected, in A549 cells (*KRAS* ^G12S^) and CFPAC cells (*KRAS* ^G12V^), the effect of sotrasib/adagrasib was almost negligible, but combination therapy with 30 nM trametinib and 1 μM imatinib showed a high inhibitory effect (Fig. 2e). On the other hand, in *KRAS* G12C mutant H23 cells and MiaPaCa-2 cells, combination therapy with 30 nM trametinib and 1 μM imatinib showed a proliferation inhibitory effect comparable to that of 0.1∼1 μM sotrasib or 0.1∼1 μM adagrasib monotherapy (Fig. 2f).

Together, these results indicate that in *KRAS*-mutated cancers, compensatory tyrosine kinase activation by trametinib and compensatory MEK/ERK activation by imatinib are resistance mechanisms to trametinib and imatinib monotherapy, respectively, and that combination therapy with trametinib and imatinib produces a synergistic antitumor effect by inhibiting these compensatory activations (Extended Data Fig. 3c). Furthermore, we found that the combination of trametinib and imatinib suppresses proliferation and survival of various *KRAS* mutant cells, highlighting that this combination therapy is a potential therapeutic approach for cancers with various *KRAS* mutations other than the G12C mutation.

### Combined imatinib and trametinib treatment induces ferritinophagy and ferroptosis

The effective combination therapy shown above prompted us to investigate its underlying mechanism in *KRAS*-mutated cancer cells. To this end, we performed RNA sequencing of *KRAS* mutant cells treated or untreated with imatinib plus trametinib and compared their transcriptomic profiling. Pathway analysis of the DEGs revealed enrichment of macroautophagy and oxidative stress-related pathways, with prominent activation of autophagosome assembly and autophagosome organization (Fig. 3a and Extended Data Fig. 4a). To further investigate which genes are involved, we analyzed the DEGs and identified increased expression of genes such as the autophagy regulators LC3-I/II (*MAP1LC3A* and *MAP1LC3B*), the ferritinophagy adaptor NCOA4, and the oxidative stress transcription factor HIF-1α, following combination treatment (Extended Data Fig. 4b). Western blotting further validated the induction of LC3-I/II, NCOA4 and HIF-1α expression in *KRAS* mutant A549 and CFPAC cells treated with imatinib plus trametinib, whereas the induction was not observed in *KRAS* WT H1975 or HCC827 cells (Fig. 3b).

**Fig. 3:**
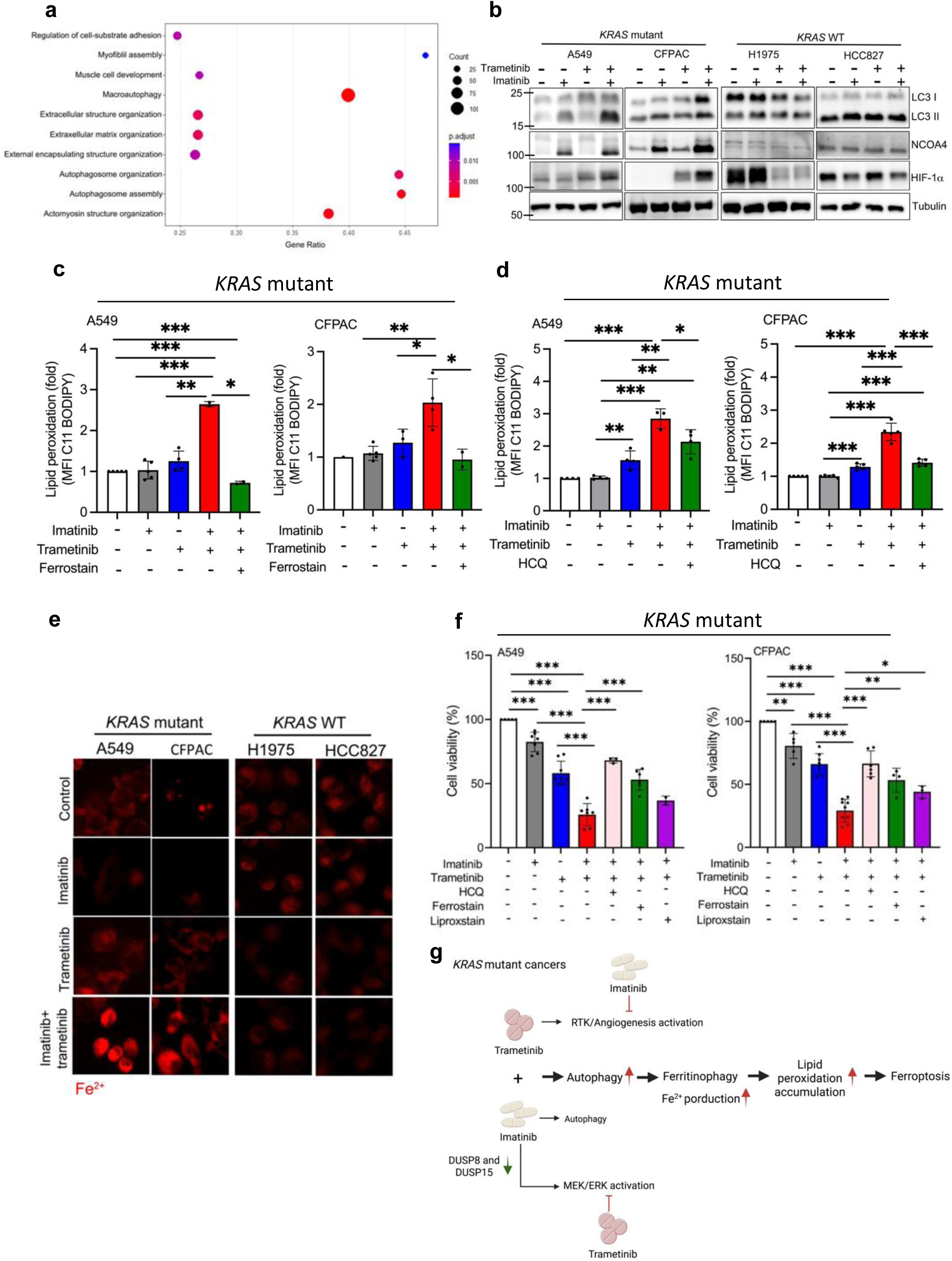
Combined imatinib and trametinib treatment induces ferritinophagy and ferroptosis. **a**, Dot plot of pathway analysis of the genes up-regulated in *KRAS* mutant cells treated with the combination of imatinib and trametinib compared to untreated cells using KEGG datasets. **b,** Western blot analysis of LC3I/II, NCOA4 and HIF-1α in A549, CFPAC, H1975 and HCC827 cells treated with imatinib and/or trametinib for 72 h. **c,** The levels of lipid peroxidation in A549 and CFPAC cells treated with 1 µM imatinib and/or 30 nM trametinib in the presence or absence of 1 µM ferrostatin for 72 h. Data represent mean ± SD from at least three independent experiments. *p < 0.05, **p < 0.01, ***p < 0.001. **d,** The levels of lipid peroxidation in A549 and CFPAC cells treated with 1µM imatinib and/or 30nM trametinib in the presence or absence of 10µM hydroxychloroquine (HCQ) for 72 h. Data represent mean ± SD from at least three independent experiments. *p < 0.05, **p < 0.01, ***p < 0.001. **e,** Confocal microscopy analysis of Fe²⁺ using iron sensing dye FerroOrange in A549, CFPAC, H1975, and HCC827 cells treated with 1µM imatinib, 30 nM trametinib, or the combination for 72 h. **f,** Colony formation assay of A549 and CFPAC cells treated with 1 µM imatinib and/or 30 nM trametinib in the presence/absence of 10 µM HCQ, 1 µM ferrostatin, or 2 µM liproxstatin for 6 days. Data represent mean ± SD from at least three independent experiments. *p < 0.05, **p < 0.01, ***p < 0.001. **g,** Graphical summary of the mechanism of ferritinophagy and ferroptosis induction by the combination of imatinib and trametinib in *KRAS* mutant cells.

The simultaneous upregulation of LC3-I/II, NCOA4, and HIF-1α demonstrates that this drug combination elicits a metabolic stress response, which triggers robust, NCOA4-dependent ferritin degradation (ferritinophagy), thereby disrupting intracellular iron homeostasis and subsequently executing ferroptosis via massive reactive oxygen species (ROS) accumulation. Therefore, we next evaluated lipid peroxidation, a characteristic of ferroptosis, using C11-BODIPY staining. The combination treatment markedly increased lipid peroxidation in *KRAS* mutant A549 and CFPAC cells, but not in *KRAS* WT cells, and this effect was abrogated by ferroptosis inhibitor ferrostatin (Fig. 3c and Extended Data Fig. 4c). Confocal imaging also demonstrated increased lipid peroxidation following combination treatment, which was reversed by ferrostatin (Extended Data Fig. 4d). In addition, the effect was also abrogated by autophagy inhibitor hydroxychloroquine (HCQ) in *KRAS* mutant A549 and CFPAC cells, suggesting that autophagy induction is the upstream event of lipid peroxidation accumulation (Fig. 3d). Because ferroptosis is driven by iron-dependent lipid peroxidation, intracellular Fe²⁺ levels were measured using the iron-staining dye, FerroOrange. Imatinib plus trametinib significantly increased labile Fe²⁺ in the *KRAS* mutant cells, whereas no increase was detected in the *KRAS* WT cells (Fig. 3e).

To further determine the functional consequence of ferroptosis induction, clonogenic survival assays were performed. Combined imatinib and trametinib treatment significantly reduced colony formation in *KRAS* mutant cells compared with the single agent treatment or control. The reduction was partially rescued by inhibition of autophagy with HCQ or ferroptosis inhibitors, ferrostatin or liproxstatin only in *KRAS* mutant but not *KRAS* WT cells, indicating that autophagy-dependent ferroptosis contributes to reduction of clonogenic survival (Fig. 3f and Extended Data Fig. 4e).

Together, these findings suggest that combination therapy with trametinib and imatinib reprograms metabolism and strongly induces autophagy, thereby promoting Fe²⁺ accumulation via ferritinophagy, which contributes to lipid peroxidation accumulation and ultimately induces cell death in *KRAS*-mutated cancers. (Fig. 3g).

### Combined imatinib and trametinib treatment suppresses KRAS-mutant tumors growth in pancreatic and lung cancer orthotopic xenograft models

Next, we investigated the therapeutic efficacy of imatinib plus trametinib *in vivo*. First, we used a PDAC orthotopic xenograft model, in which *KRAS* G12V mutant CFPAC human PDAC cells were orthotopically inoculated in NPG mice. Subsequently, the therapeutic effects of imatinib, the trametinib, and the imatinib and trametinib combination were evaluated. In all the mouse models used here, mice were orally administered inhibitors equivalent to clinically relevant human exposure levels daily for 30 days. Monotherapy with either imatinib or trametinib did not significantly affect tumor growth, whereas the combination treatment markedly suppressed tumor progression (Fig. 4a, b and Extended Data Fig. 5a). The survival time of mice was significantly extended beyond 30 days in the combination treatment group compared with the untreated control group or the groups receiving imatinib or trametinib monotherapy (Fig. 4c). Since the toxicity of the combination therapy is a major concern in clinical setting, the body weight of these mice was also monitored every 3 days. There was no significant effect on body weight in any of the treatment groups including combination therapy (Extended Data Fig. 5b). During the treatment course, mice receiving trametinib monotherapy exhibited significantly higher incidence of ascites formation and spontaneous mortality compared with other groups. This observation may partially explain the limited clinical efficacy of trametinib reported in previous clinical trials ^38–40^. Moreover, these findings support the premise that combining trametinib with imatinib may enhance its therapeutic efficacy with no sign of increased toxicity.

**Fig. 4:**
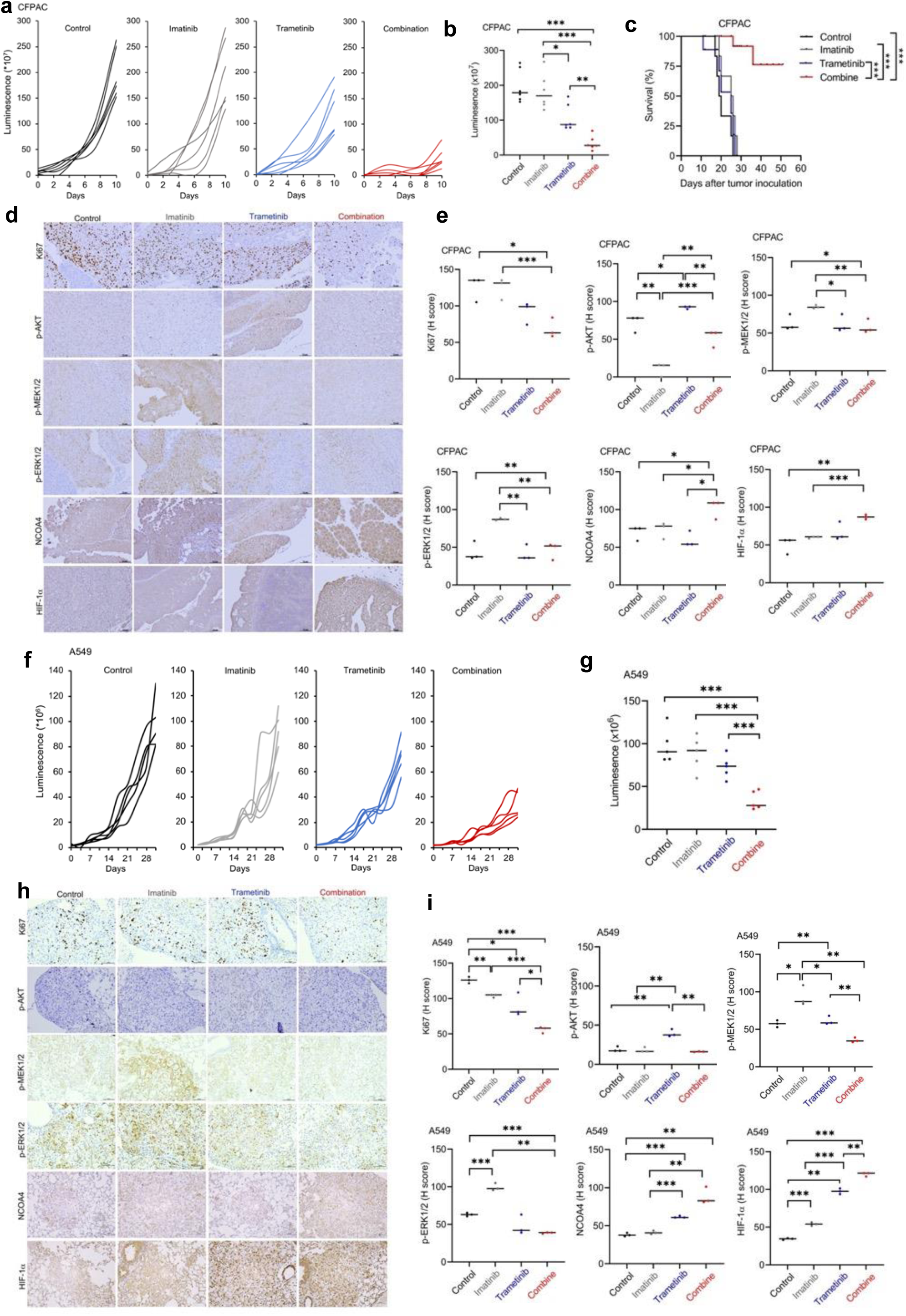
Combined imatinib and trametinib treatment suppresses *KRAS* mutant tumors growth. **a**, Tumor growth curves of NPG mice bearing CFPAC *KRAS* G12V mutant pancreatic cancer xenografts treated with vehicle, imatinib (15 mg/kg), trametinib (0.3 mg/kg), or the combination. Data represent mean ± SEM; n = 6 mice/group. **b,** Luminescence signals representing xenograft tumor sizes on day 30 used in (**a**) (n=6 mice/group). Data represent mean ± SD from six mice. *p < 0.05, **p < 0.01, ***p < 0.001. **c,** Survival curve of the mice used in (**a**). **d,** Representative IHC images (20x) of Ki67, p-AKT (S473), p-MEK1/2 (S217/221), p-ERK1/2 (Y204/T202), NCOA4 and HIF-1⍺ in the tumors of the mice on day 30 used in (**a**). **e,** Quantification of IHC staining shown in (**d**). Data represent mean ± SD from three mice. *p < 0.05, **p < 0.01, ***p < 0.001. **f,** Tumor growth curves of ASID mice bearing A549 *KRAS* G12S mutant lung cancer xenografts treated orally with vehicle, imatinib (15 mg/kg), trametinib (0.3 mg/kg), or the combination. Data represent mean ± SEM; n = 5 mice/group. **g,** Luminescence signals representing xenograft tumor sizes on day 30 used in (**f**) (n=5 mice/group). Data represent mean ± SD from five mice. ***p < 0.001. **h,** Representative IHC images (20x) of Ki67, p-AKT (S473), p-MEK1/2 (S217/221), p-ERK1/2 (Y204/T202), NCOA4 and HIF-1⍺ in the tumors of the mice on day 30 used in (**f**). **i,** Quantification of IHC staining shown in (**h**). Data represent mean ± SD from three mice. *p < 0.05, **p < 0.01, ***p < 0.001.

We next analyzed signaling alterations in tumor tissues by IHC. The results showed the marked inhibition of the proliferation marker Ki67 and p-AKT, p-MEK1/2, and p-ERK1/2, along with increased expression of ferritinophagy-related markers, NCOA4 and HIF-1α in the tumors treated with the combination of imatinib and trametinib (Fig. 4d, e). Imatinib monotherapy increased p-MEK1/2 and p-ERK1/2 in *KRAS* mutant tumors, highlighting feedback activation of the MEK/ERK pathway as observed in the *vitro* study. In contrast, trametinib monotherapy increased p-AKT in *KRAS* mutant tumors. Consistent with this observation, trametinib alone enhanced expression of the micro-vessel marker, CD31 expression (Fig. 1e, g), suggesting that the feedback activation of these RTK signaling and angiogenesis may contribute to limitation of trametinib monotherapy in *KRAS* mutant tumors. The antitumor activity was further confirmed by Hematoxylin and Eosin (H&E) staining (Extended Data Fig. 5c).

Although *KRAS* G12C mutation is the most common *KRAS* mutation in NSCLC, more than half of NSCLC with *KRAS* mutation has non-G12C mutation. Therefore, we evaluated the therapeutic efficacy of imatinib plus trametinib in an orthotopic lung cancer xenograft model using ASID mice bearing *KRAS* G12S mutant A549 human NSCLC cells. Notably, similar results were observed in the *KRAS* G12S mutant NSCLC model as in the PDAC model described above. Combination treatment significantly suppressed tumor progression, whereas imatinib or trametinib monotherapy did not affect tumor growth (Fig. 4f, g). Body weight remained unchanged across all treatment groups, suggesting acceptable tolerability (Extended Data Fig. 5d). IHC analyses also revealed reduced expression of Ki67 and p-AKT, p-MEK1/2, and p-ERK1/2, along with increased expression of ferritinophagy markers NCOA4 and HIF-1α in tumors treated with the combination of imatinib and trametinib (Fig. 4h, i). Moreover, antitumor activity was further supported by H&E staining (Extended Data Fig. 5e).

Finally, we evaluated liver and kidney toxicity in the A549 orthotopic xenograft model described in Fig 4f-i. Serum markers of liver and kidney function, including alanine aminotransferase (ALT), aspartate aminotransferase (AST), creatinine, and blood urea nitrogen (BUN), remained within normal ranges in treated mice (Extended Data Fig. 5f). Thus, the combination treatment maintained robust antitumor activity over one month without evidence of systemic toxicity.

Taken together, these findings reveal that monotherapy with imatinib or trametinib activates compensatory survival signals and that the combination of these two drugs suppresses their respective compensatory survival signals *in vivo*. Thus, the combination of imatinib and trametinib effectively suppresses the growth of *KRAS*-mutated tumors, at least part, through the induction of ferritinophagy and ferroptosis.

### The effects of combined imatinib and trametinib treatment in primary pancreatic cancer cells and PDAC patient-derived xenograft tumors

To further investigate and extend the clinical potential of the imatinib plus trametinib combination, we validated the results obtained so far using primary *KRAS* G12D mutant PDAC cells isolated from a PDAC patient and it’s PDX model and obtained similar results. In brief, we first examined p-AKT, p-ERK1/2, LC3I/II, NCOA4, and HIF-1α expression in the primary pancreatic cancer cells after 3 days of treatment with vehicle, imatinib or trametinib monotherapy, or the combination of imatinib and trametinib. Similar to the results obtained from *KRAS*-mutated cell lines (Figs. 2-3), the LC3I/II, NCOA4, and HIF-1α expression was increased in the combination treatment group compared with untreated or monotherapy groups (Fig. 5a). We next evaluated the therapeutic efficacy of the combination of imatinib and trametinib in a *KRAS* G12D mutant PDAC PDX model (Fig. 5b). Consistently, the combination of imatinib and trametinib significantly delayed tumor growth and extended median survival without affecting body weight compared to the untreated control and monotherapy (Fig. 5c-e, and Extended Data Fig. 6a). The data also demonstrated suppression of Ki67 and p-AKT, p-MEK1/2, and p-ERK1/2 accompanied by induction of ferritinophagy markers NCOA4 and HIF-1α in tumors treated with the combination of trametinib and imatinib (Fig. 5f-h). And imatinib monotherapy increased p-MEK1/2 and p-ERK1/2, while trametinib monotherapy enhanced p-AKT in the PDX tumors. The antitumor activity was also supported by H&E staining (Extended Data Fig. 6b).

**Fig. 5:**
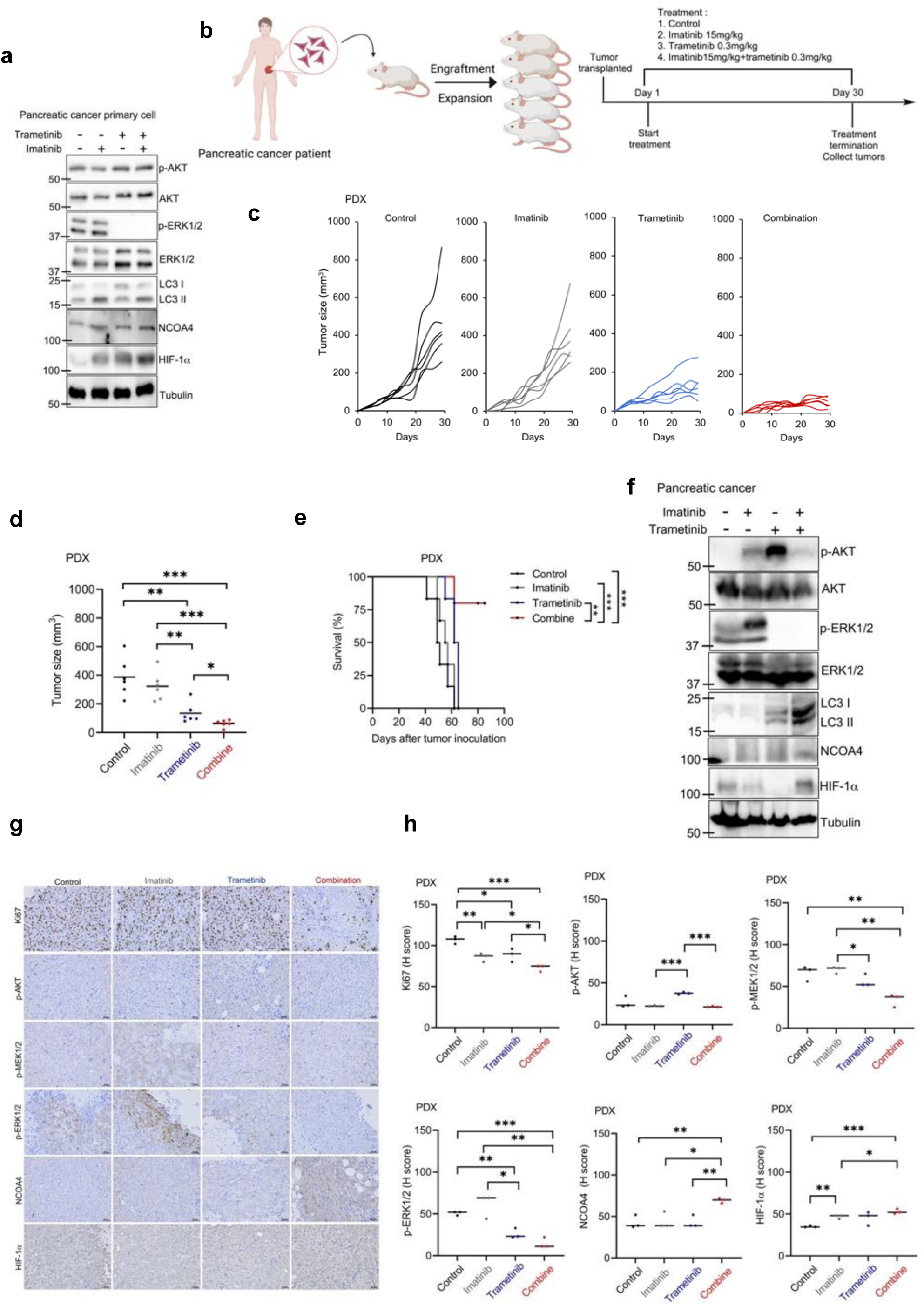
The effects of combined imatinib and trametinib treatment in primary pancreatic cancer cells and pancreatic cancer patient-derived xenograft tumors. **a**, Primary *KRAS* G12D mutant pancreatic cancer cells were treated with imatinib, trametinib, or the combination at the indicated concentrations for 72 h, and their lysates were analyzed by western blot to detect p-AKT (S473), AKT, p-ERK1/2(Y204/T202), ERK1/2, LC3I/II, NCOA4 and HIF-1⍺ expression. **b,** Schematic diagram of generation of pancreatic patient-derived xenograft (PDX) mice. **c,** PDX tumor growth curves of NPG mice treated with vehicle, imatinib (15 mg/kg), trametinib (0.3 mg/kg), or the combination. Data represent mean ± SEM; n = 6 mice/group. **d,** Tumor size of the mice on day 30 used in (**c**). Data represent mean ± SD from six mice. *p < 0.05, **p < 0.01, ***p < 0.001. **e,** Survival curve of the mice used in (**c**). **f,** Western blot analysis of p-AKT (S473), AKT, p-ERK1/2 (Y204/T202), ERK1/2, LC3I/II, ferroptoisis-associated genes, including NCOA4 and HIF-1⍺ in the tumors of the mice on day 30 used in (**c**). **g,** Representative IHC images (20×) of Ki67, p-AKT (S473), p-MEK1/2 (S217/221), p-ERK1/2 (Y204/T202), NCOA4 and HIF-1⍺ in the tumors of the mice on day 30 used in (**c**). **h,** Quantification of IHC staining shown in (**g**). Data represent mean ± SD from three mice. *p < 0.05, **p < 0.01, ***p < 0.001.

Taken together, the findings from the mouse models including orthotopic xenograft and PDX models demonstrate that combination therapy with imatinib and trametinib is a powerful therapeutic approach for *KRAS*-mutated cancers.

### The combination of trametinib and Imatinib also suppresses KRAS Inhibitor-resistant NSCLC cells

The development and clinical implementation of KRAS G12C inhibitors represents one of the recent breakthroughs in cancer therapy. However, the emergence of resistance remains a significant clinical challenge. Therefore, we asked whether the combination of trametinib and imatinib could overcome resistance in sotorasib-resistant *KRAS* G12C-mutated NSCLC cells. To this end, we first developed a sotorasib-resistant line, H23R from the H23 NSCLC line carrying *KRAS* G12C mutation. We treated parental H23 cells and sotorasib-resistant H23R cells with different concentrations of sotorasib and examined their cell viability, and confirmed that H23R cells maintained higher viability than parental H23 cells, indicating resistance to sotorasib (Fig. 6a). Next, we compared the signaling profiles of sotorasib-resistant H23R cells with their parental H23 cells (Fig. 6b, c). Western blot analysis revealed selectively increased phosphotyrosine levels accompanied by AKT activation in the resistant H23R cells, as comparing with the sensitive parental cells (Fig. 6b). Furthermore, we detected increased p-EGFR and p-PDGFRα/β, in the H23R cells (Fig. 6c). These data suggest that the activation of multiple RTKs and the AKT pathway may contribute to the sotorasib resistance in H23R cells.

**Fig. 6:**
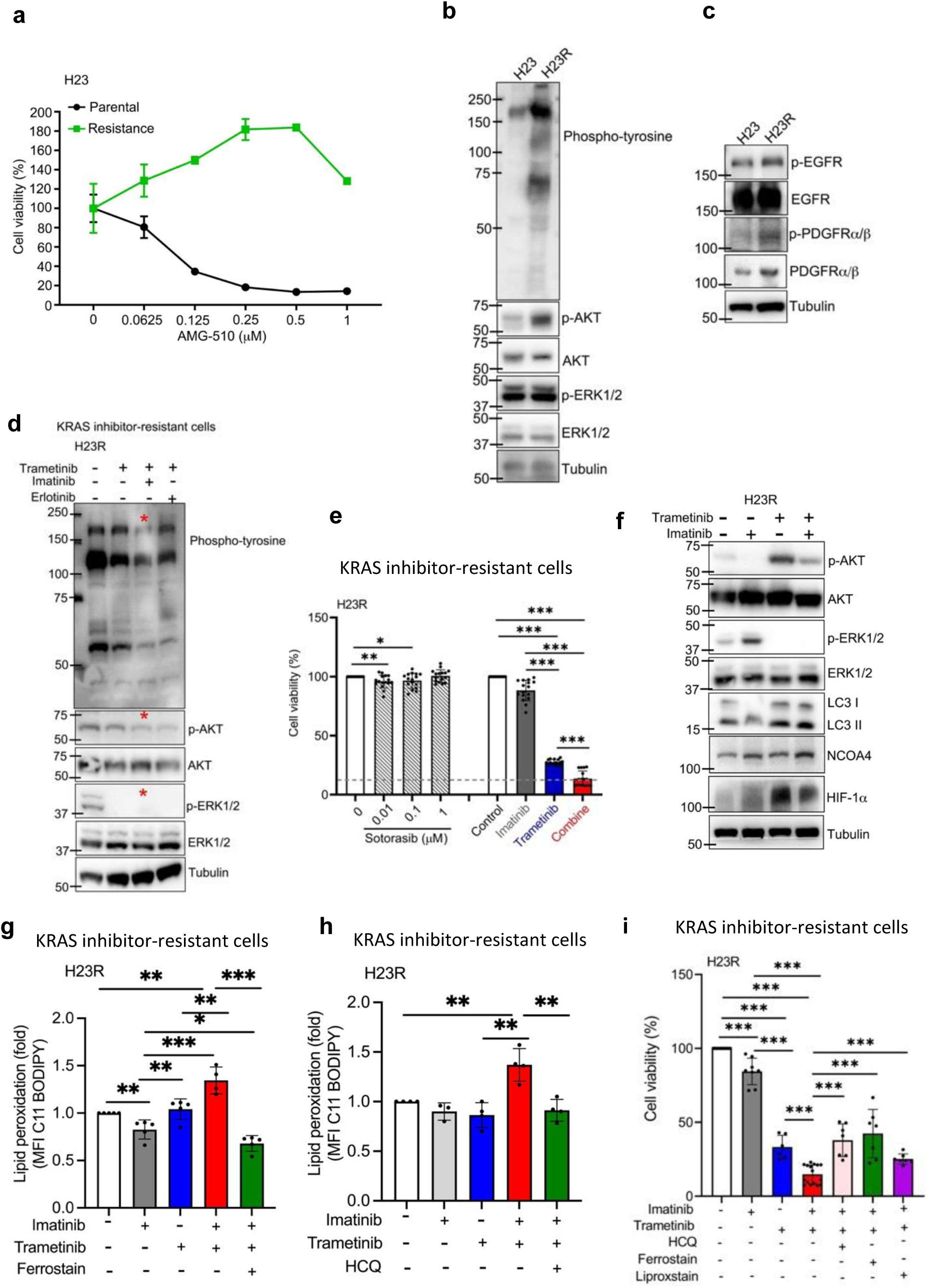
Trametinib-Imatinib combination suppresses growth and signaling in KRAS Inhibitor resistant NSCLC cells. **a**, Cell viability assay comparing parental H23 and sotorasib (AMG-510)-resistant H23R cells treated with the indicated concentrations of sotorasib. Data are presented as mean ± SD. **b,** Western blot analysis of phosphorylated tyrosine kinases, p-AKT (S473), AKT, p-ERK1/2 (Y204/T202), and ERK1/2 in H23 and H23R cells. **c,** Western blot analysis of p-EGFR (Y1068), EGFR, p-PDGFR⍺ (Y849) /PDGFRβ (Y857) and PDGFR⍺/β in H23 and H23R cells. **d,** Western blot analysis of phosphorylated tyrosine kinases, p-AKT (S473), AKT, p-ERK1/2 (Y204/T202), and ERK1/2 in H23R cells following treatment with trametinib alone or in combination with imatinib (1 μM) or erlotinib (1 μM). **e,** Colony formation assays of H23R cells treated with sotorasib, imatinib (1 μM), trametinib (30 nM), or the imatinib-trametinib combination at the indicated concentrations for 6 days. Data represent mean ± SD from at least three independent experiments. *p < 0.05, **p < 0.01,***p < 0.001. **f,** Western blot analysis of p-AKT (S473), AKT, p-ERK1/2 (Y204/T202), ERK1/2, LC3I/II, NCOA4 and HIF-1α in H23R cells treated with imatinib and trametinib for 24 h. **g,** The levels of lipid peroxidation in H23R cells treated with imatinib (1 μM) and/or trametinib (30 nM), in the presence/absence of ferrostatin (1 μM) for 2 days. Data represent mean ± SD from at least three independent experiments. *p < 0.05, **p < 0.01, ***p < 0.001. **h,** The levels of lipid peroxidation in H23R cells treated with imatinib (1 μM) and/or trametinib (30 nM), in the presence/absence of hydroxychloroquine (HCQ) 10 μM for 2 days. Data represent mean ± SD from at least three independent experiments. **p < 0.01. **i,** Colony formation assay of H23R cells treated with imatinib (1 μM) and/or trametinib (30 nM) in the presence/absence of hydroxychloroquine (HCQ, 10 μM), ferrostatin (1 μM), or liproxstatin (2 μM) for 6 days. Data represent mean ± SD from at least three independent experiments. ***p < 0.001.

Consistent to the results from Fig. 2b, the combination of trametinib and imatinib, but not trametinib with erlotinib, effectively reduced phosphotyrosine signaling (Fig. 6d). Colony formation assays demonstrated that the combination of trametinib and imatinib suppressed clonogenic survival more potently than either monotherapy or sotorasib in H23R cells (Fig. 6e). Western blotting analysis indicated that the combination therapy induced the expression of LC3 I/II, NCOA4 and HIF-1α in H23R cells (Fig. 6f). Increased lipid peroxidation was observed in H23R cells, and this effect was abrogated by ferroptosis inhibitor ferrostatin (Fig. 6g). In addition, the effect was also suppressed by autophagy inhibitor HCQ in H23R cells (Fig. 6h).

Next, to determine the functional consequence of ferroptosis induction, clonogenic survival assays were conducted. Combined treatment with imatinib and trametinib significantly reduced colony formation in sotorasib-resistant H23R cells compared with single agent treatment or control. The reduction was partially rescued by inhibition of autophagy with HCQ or by ferroptosis inhibitors, ferrostatin or liproxstatin, suggesting that autophagy-dependent ferroptosis contributes to loss of clonogenic survival of H23R cells (Fig. 6i).

Taken together, these results suggest that the combination of trametinib and imatinib also induces ferroptosis in KRAS inhibitor-resistant cells. Consequently, this combination therapy may represent a promising strategy to overcome resistance to KRAS G12C inhibitors.

### Evaluation of efficacy and toxicity of the combination of trametinib and imatinib in pancreatic cancer patients

On the basis of these preclinical findings, we initiated a compassionate-use, pilot study to evaluate the combination therapy of trametinib plus imatinib in patients with chemotherapy-refractory pancreatic adenocarcinoma harboring non-*KRAS* G12C mutations at China Medical University Hospital (NCT06962254). Eligible participants had progressive disease after at least two prior lines of standard therapy and their tumors had non-*KRAS* G12C mutations.

As of the data cutoff on May 4, 2026, four participants, two with *KRAS* G12R mutation, one with *KRAS* G12V mutation, and one with *KRAS* G12D mutation, had received the combination therapy (Extended Data Fig. 7a, b). Three participants (75%) are females, and the median age is 68 years, with a range from 63 to 75 years. Three of four participants showed biochemical response or control of serum carbohydrate antigen 19-9 (CA19-9) (Fig. 7a) within 8-16 weeks of treatment. Although none had complete or partial response, the disease control rate (DCR) was 75% (Extended Data Fig. 7c). The median PFS and overall survival (OS) was 8 weeks (95% CI:0 to 15.48) and 21.57 weeks (95 % CI: 3.6 to 39.53), respectively (Fig. 7b, c).

**Fig. 7:**
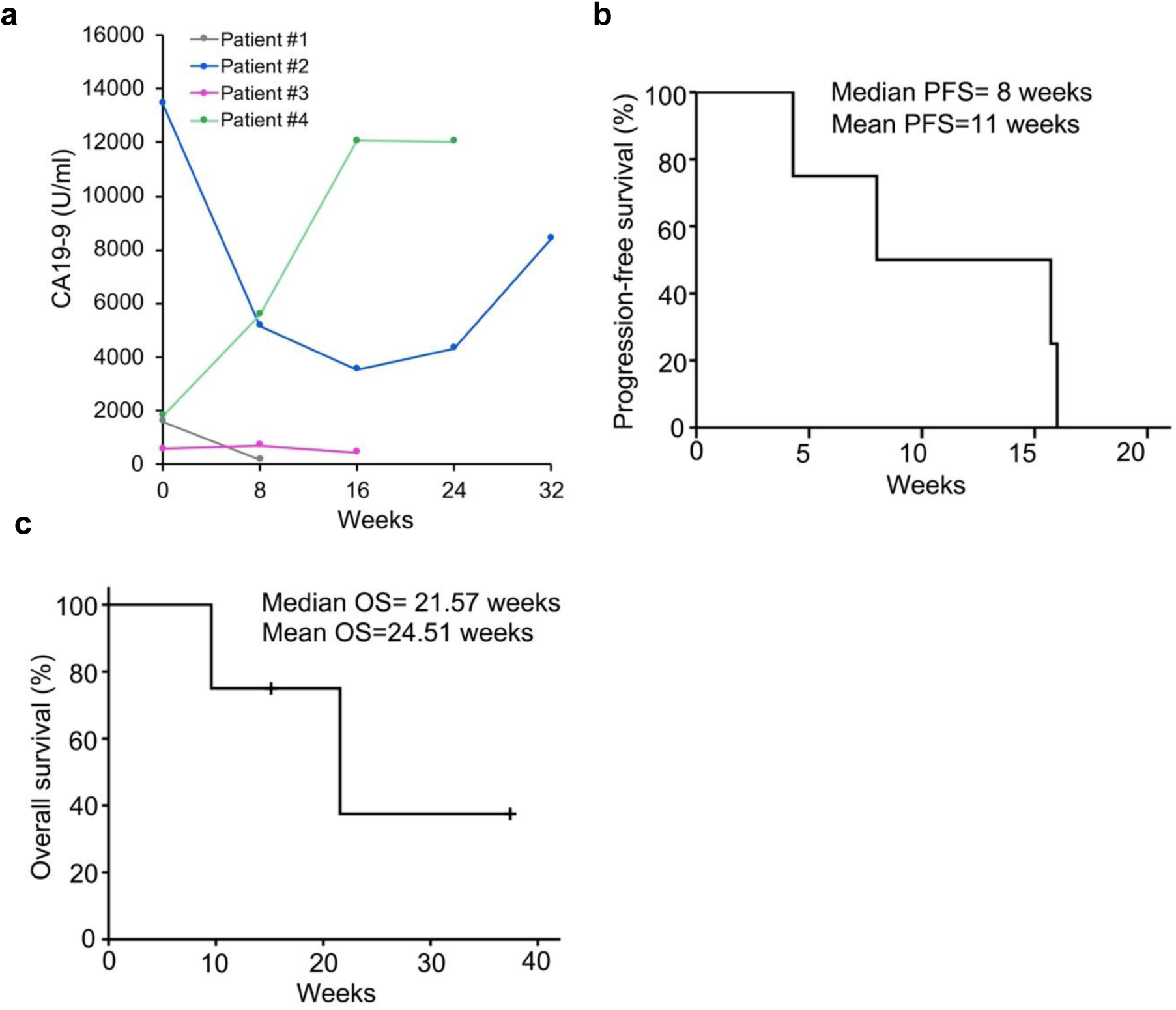
Clinical evaluation of the combination of imatinib and trametinib in patient with *KRAS* non-G12C mutant, chemotherapy-refractory pancreatic cancer a, Change of carbohydrate antigen 19-9 (CA19-9) of four participants following start of therapy. **b,** Kaplan-Meier plots of progression-free survival (PFS). **c,** Kaplan-Meier plots of overall survival (OS).

The observed treatment-emergent adverse events included increased AST, increased ALT, diarrhea, rash, abdominal pain, mucositis and erythematous papules, with all were grade 1 in degree (Table 1). Thus, this pilot trial with limited number of patients suggest the combination therapy is relatively safe and reaches stable disease with extended survival rate.

**Table 1.** Treatment emergent adverse event for all participants (N = 4).

| | Grade 1 | Grade 2 | Grade $\geq 3$ |
| --- | --- | --- | --- |
| Increased aspartate aminotransferase (AST), n | 2 | 0 | 0 |
| Increased alanine aminotransferase (ALT), n | 2 | 0 | 0 |
| Diarrhea, n | 1 | 0 | 0 |
| Rash, n | 1 | 0 | 0 |
| Abdominal pain, n | 1 | 0 | 0 |
| Mucositis, n | 1 | 0 | 0 |
| Erythematous papules, n | 1 | 0 | 0 |

## Discussion

This study provides a rationale for a novel combination strategy that simultaneously targets the MEK and RTKs pathways for the treatment of *KRAS* mutant cancers, including those resistant to FDA-approved KRAS inhibitors. *KRAS*-driven malignancies had been considered undruggable due to the diversity of oncogenic mutations, such as G12C, G12D, G12V, G12S and Q61H, as well as intrinsic structural constraints of the KRAS protein. The recent clinical approval of sotorasib and adagrasib, which selectively target *KRAS* G12C, represents a significant advance in this field. However, the clinical efficacy remains limited by mutation specificity and the rapid emergence of adaptive resistance. In parallel, several pieces of evidence have highlighted substantial progress in the development of small-molecule KRAS inhibitors and in understanding the mechanisms underlying RAS pathway inhibition ^15,31,41–43^. Notably, emerging clinical studies have identified promising therapeutic candidates that specifically target *KRAS* G12D mutant cancers, and most recent data on a pan-KRAS inhibitor ^44,45^, thereby expanding the scope of KRAS-directed therapies beyond G12C ^11,13,14^. In this evolving landscape, our findings support the concept that co-inhibition of MEK and RTK signaling can not only suppress various *KRAS*-mutated cancers but also overcome some resistance to KRAS inhibitors. Collectively, these results underscore the therapeutic potential of rational combination strategies to broaden treatment efficacy and achieve more durable responses across diverse *KRAS* mutant contexts.

Extensive efforts have focused on targeting *KRAS*-driven tumors through suppression of downstream signaling using MEK inhibitors. Trametinib, a clinically approved MEK inhibitor, has demonstrated efficacy in patients with *BRAF* V600E mutant NSCLC and melanoma; however, its therapeutic benefit in *KRAS* mutant cancers remains limited ^46–49^. Accumulating evidence indicates that MEK inhibitor monotherapy frequently induces feedback activation of bypass signaling pathways, including EGFR, PI3K/AKT, FGFR pathways, thereby attenuating its antitumor efficacy ^22,50^. Consequently, the development of effective MEK-based combination strategies for *KRAS*-mutated cancers remains a significant clinical challenge. To further elucidate the mechanism underlying adaptive reactivation of survival signaling in *KRAS* mutant cancers following MEK inhibition, we investigated the molecular response to trametinib treatment. Our results reveal simultaneous activation of multiple RTKs, such as EGFR and PDGFR α/β, along with enhanced angiogenesis-associated signaling, which may collectively contribute to adaptive resistance. These findings provide mechanistic insight into the limitation of MEK inhibitor monotherapy and support the rationale for combination approaches targeting both MEK and RTK pathways. Additionally, we observed consistent activation of RTKs in both in *in vivo* and *in vitro* models following MEK inhibition, further supporting the central role of RTK-mediated pathways in the adaptive resistance. These findings promoted us to explore combinatorial strategies targeting both MEK and RTK signaling as a potential therapeutic approach for *KRAS* mutant cancers. To address this issue, we performed a screening of FDA-approved inhibitors and identified imatinib as a candidate drug. Imatinib has been shown to exert antitumor activity through inhibition of PDGFR α/β, and AKT signaling pathways ^51–53^, thereby providing a rationale for suppressing trametinib induced adaptive reactivation of survival signaling. Our findings indicated that the combination of trametinib plus imatinib significantly inhibits colony formation across a spectrum of *KRAS* mutant cancers. Consistently, this combination strategy markedly suppressed tumor growth in multiple *KRAS* mutant xenograft models, including *KRAS* G12V PDAC and *KRAS* G12S NSCLC xenograft models as well as a *KRAS* G12D PDAC PDX model.

Mechanistically, the combination of trametinib and imatinib effectively suppresses both the RTK/AKT and MEK/ERK signaling pathways in *KRAS-*mutated cancers, resulting in metabolic reprogramming and robust autophagy, leading to ferritinophagy and ferroptosis in cancer cells. This may be the mechanism of the observed synergistic antitumor effects. Notably, imatinib inhibits the RTK-driven adaptive signaling, thereby preventing the reactivation of AKT and angiogenic pathways induced by prolonged trametinib exposure. On the other hand, trametinib can suppress MEK/ERK activation induced by imatinib treatment. Thus, the simultaneous inhibition of these pathways effectively suppresses both adaptive feedback-signaling and reinforces pathway blockade, thereby enhancing therapeutic efficacy. This mechanistic insight provides a strong rationale for clinical translation of trametinib combined with imatinib as an effective alternative therapeutic strategy for *KRAS* mutant cancers.

The clinical translation of mechanistic insights remains a critical determinant of their therapeutic impact. Although multiple preclinical studies have proposed combination strategies, such as MEK inhibitors with chemotherapy or with SHP1 and SOS1 inhibitors, these approaches have far failed to demonstrate substantial clinical benefit in patients harboring the *KRAS* mutations ^38^. Notably, the successful application of trametinib based combination with *BRAF* V600E mutant NSCLC has renewed confidence in the development of rational combinatorial approaches targeting oncogenic signaling pathways. In this context, our study provides preliminary clinical evidence supporting the therapeutic potential of trametinib combined with the multi-tyrosine kinase inhibitor imatinib. In the pilot study of four participants with advanced PDAC harboring various non-G12C *KRAS* mutations, this combination regimen resulted in sustained control of tumor burden in target lesions. Although limited by tumor size, these findings suggest potential clinical activity in a patient population with otherwise limited treatment options. Importantly, safety remains a key consideration in early-phase clinical trial. Our results indicate that combination of trametinib and imatinib was generally well tolerated, with no grade ≥ 2 advance events (AEs) observed. The most common treatment-related AEs were grade 1 toxicity, including rash, diarrhea, hepatic injury. These observations support a manageable safety profile this combination regimen. Taken together, the observed clinical activity and favorable tolerability highlight the potential of trametinib plus imatinib as a promising therapeutic strategy for patients with non-G12C *KRAS* mutant PDAC.

Moreover, considering cost-effectiveness, the combination therapy of imatinib and trametinib has extremely high health economic value. Imatinib’s patent expired more than 10 years ago, and low-cost generic versions are now widely available. Therefore, the cost of imatinib would be less than 200 US dollar per month. Trametinib is still under patent protection and is around $5,000 to $10,000 per month, but generic versions are expected to enter the market within the next 1 to 3 years. In contrast, the cost of KRAS inhibitors exceeds $20,000 per month, and generic versions are not expected over the next 12 years ^54^. Thus, assuming long-term treatment continuation, this combination therapy has a significant economic advantage over KRAS inhibitors.

Overall, the combination of trametinib plus imatinib demonstrated promising therapeutics efficacy. However, the study has several limitations. First, the small size of this pilot-IIT, which included only four participants with non-G12C *KRAS* mutant PDAC, limits the generalizability of the findings. Therefore, further validation in larger, randomized controlled trials is warranted to establish the clinical efficacy and safety of this combination regimen. Moreover, because the combination of trametinib and imatinib has a potential to overcome the resistance to KRAS-G12C inhibitors, the validation in NSCLC patients resistant to KRAS-G12C inhibitors would also be necessary. Second, the identification of predictive biomarkers for patient selection remains a critical unmet need. Interestingly, the resistant cell lines used in this study showed enhanced tyrosine phosphorylation signaling (Fig 6b, c). Considering that diverse mechanisms of resistance to KRAS G12C inhibitors exist, the activity status of tyrosine phosphorylation signaling in cancer tissues would serve as a predictive biomarker for this combination therapy.

In summary, this study represents a comprehensive translation effort, spanning from preclinical investigation to early-phase clinical evaluation, and highlight the therapeutic potential of co-targeting MEK/RTK pathways in patients with *KRAS* mutant PDAC. These findings provide a strong rationale for further clinical development of this combinatorial strategy.

## Material and method

### Cell line and culture

All cell lines, including human NSCLC cell lines A549 (*KRAS* ^G12S^), H441 (*KRAS* ^G12V^), H23(*KRAS* ^G12C^), H1975(*KRAS* ^WT^), and HCC827(*KRAS* ^WT^), as well as human PDAC cell lines CFPAC (*KRAS* ^G12V^), MiaPaCa 2 (*KRAS* ^G12C^), and SU.86.86 (*KRAS* ^G12D^) were obtained from the American Type Culture Collection (ATCC). The cells were maintained in a humidified incubator at 37 °C with 5% CO_2_ and grown in RPMI1640 or Dulbecco’s Modified Eagle Medium (DMEM) supplemented with 10% fetal bovine serum (FBS) and penicillin (100U/ml)/streptomycin (100µg/ml) (Gibco/Thermo Fisher). All cell lines have been tested for microplasma contamination and were validated by short tandem repeat (STR) DNA fingerprinting the AmpFLSTR® PCR Amplification Kit (Life Technologies). The STR profiles were compared with ATCC fingerprints and the cell line integrated molecular authentication database. H23 sotorasib-resistance cells (H23R) was generated by continuous (>2 months) culture in standard RPMI1640 medium in the presence of 1 µM sotorasib.

### Reagents and antibodies

Imatinib, trametinib, and hydroxychloroquine were purchased from MedChemExpress. Ferrostatin and liproxstatin were purchased from Cayman Chemistry. All inhibitors were dissolved in dimethyl sulfoxide (DMSO) or ddH_2_O.

The primary antibodies and dilution ratios for western blot analysis were follows: Rabbit anti-EGFR (#4276, 1:1,000), rabbit anti-PDGFR (#3169, 1:1,000), rabbit anti-PDGFR α/β at Y849/Y857 (#3170, 1:1,000), rabbit anti-p-MEK1/2 S217/221 (#9154, 1:1,000), mouse anti-MEK1/2 (#4694, 1:1,000), rabbit anti-ERK1/2 Y204/T202 (#4370S, 1:1,000), rabbit anti-AKT S473 (#4060L, 1:1,000), mouse anti-AKT (#9272S, 1:1,000), rabbit anti-CRaf S338 (#9427, 1:1,000), rabbit anti-BRaf S445 (#2696, 1:1,000), rabbit YAP (#14074, 1:1,1000), rabbit TAZ (#83669, 1:1,000), rabbit cleaved PARP (#9532, 1:1,000) and rabbit anti-HIF-1α (#31619, 1:1,000) are from Cell Signaling Technology (CST); Rabbit anti-EGFR Y1068 (AP0301, 1:1,000) and DUSP15 (A16322, 1:1,000) are from Abclonal; Rabbit anti-NCOA4 (GTX32739, 1:1,000), rabbit PPP1R3C (GTX87429, 1:1,000) and mouse phospho-tyrosine (GTX14167, 1:1,000) are from GeneTex; Rabbit anti-LC3 (NB100-2220, 1:1,000) is from Novus Biologicals; Mouse anti-tubulin (sc-5286, 1:5,000), mouse anti-ERK1/2 (sc-514302, 1:1,000) and DUSP18 (sc-376923, 1:1,000) are from Santa Cruz Biotechnology. Rabbit anti-CD31 (ab28364, 1:1,000) is from Abcam. Rabbit DUSP8 (A93212, 1:1,1000) is from EBL Biotechnology; Rabbit PPP3CA (13422-1-AP, 1:1,000) is from Proteintech; Rabbit PPP1R21 (A305-825A, 1:1,000) is from Bethyl.

### Clonogenic assay

For the clonogenic assay, three thousand cells were seeded in 24-well plates in RPMI1640 or DMEM medium containing 10 %FBS overnight, then treated with respective agent(s). The medium was replaced every three days. After the indicated days, cells were washed with phosphate-buffered saline (PBS), fixed with 4% formaldehyde, and stained with 0.5% crystal violet. Crystal violet was dissolved in 5% SDS solution overnight and optical density of each well measured at OD570 nm (OD 570) using an ELISA plate reader. The average OD_570_ of control cells was set to 100%. The percentage of treated cells that were viable were then calculated accordingly. The medium inhibitory concentration (IC50) for each drug was determined from dose-effect relationship at four or five concentrations of each drug using the CompuSyn software (Compusyn, Inc.). Data are expressed as percentage of control cells and mean± s.d. of three independent experiments.

### Receptor tyrosine kinase antibody array

A Proteome Profiler Human Phospho-RTK Array Kit (R&D systems, #ARY001B) was used according to the manufacturer’s instructions. In brief, cells were treated with DMSO or 30 nM trametinib for 48 hour and then harvested for antibody array analysis. Signal data from the array were captured and analysis as Western blot images. Signals on each array were normalized to the mean signal value of reference controls.

### Flow cytometry analysis

Cells were treated/untreated with respective agent(s) for 48 hours. Then, the cells were incubated at 37°C for 20 mins in the presence/absence of the BODIPY™ 581/591 C11 (Invitrogen). After washing the cells with PBS, flow cytometric analysis was performed according to manufacturer’s instruction (CytoFlex, Beckman Coulter). Signals on each sample were normalized to the mean signal value of reference controls.

### RNA-seq library and bioinformation analysis

A549 cells were treated with 30 nM trametinib for 48 hours. Total RNA was isolated with TRIZOL solution (ThermoFisher Scientific). Assessment of RNA quality, library construction, RNA sequencing was processed and analyzed by GENEWIZ (Burlington, MA, USA). The pipeline used for RNA sequencing data analysis was Cutadapt (data cleaning), HISAT2 (alignment), and HTSeq (counting). Differentally expressed genes were analyzedc using DESeq2. The pathway analysis was performed by using g:Profiler ^55^. Gene Set Enrichment Analysis (GSEA) is performed by using GSEA software^56^ .

### Quantitative real-time PCR

Total RNA was extracted from the cells, and cDNA was synthesized by using SuperScript™ III First-Strand Synthesis kit (Invitrogen). Universal SYBR Color qPCR Master Mix (Invitrogen) was used for real-time PCR according to the manufacturer’s instructions. The expression of target genes was normalized against that of GAPDH and compared among groups via the ΔΔCT method. The sequences of primers used in this study were follows: DUSP18 forward, 5’-ATCCACAGCGTGGAGATGAAGC-3’; DUSP18 reverse, 5’-GCGTGGTACTTCATGAGGTAGG-3’; DUAP8 forward, 5’-TCATCTGCGAGAGCCGCTTCAT-3’; DUAP8 reverse, 5’-AGCCAGACAGTGGACGATGACT-3’; DUSP15 forward, 5’-TCAACTTCATCCACTGCTGCCG-3’; DUSP15 reverse, 5’-TACGCTGTCACAATCGTGGTGC-3’; PPP3CA forward, 5’-GCCCTGATGAACCAACAGTTCC-3’; PPP3CA reverse, 5’-GCAGGTGGTTCTTTGAATCGGTC-3’; PPP1R3C forward, 5’-CCTCTGCCTTAAAACACCACGAG-3’; PPP1R3C reverse, 5’-CAACGAGCAGTTCTCCAGACAG-3’; PPP1R21 forward, 5’-GACCTAAGGCAGCGAGTGGATT-3’; PPP1R21 reverse, 5’-CATAAGGCACAGACTCCAAGAGG-3’; PPP1R12A forward, 5’-GCAGGTGTTACACGTTCAGCTTC-3’; PPP1R12A reverse, 5’-GATGTACTGGCTAGTCGTCTTGG-3’.

### Western blot

Western blot analysis was performed according to standard procedures. Briefly, cells were washed twice with PBS, lysed in RIPA lysis buffer. Then, the samples were separated by Sodium dodecyl sulfate-polyacrylamide gel electrophoresis (SDS-PAGE) and transferred onto polyvinylidenes fluoride membranes (Bio-Rad). After overnight incubation with primary antibody, washing, and incubation with secondary antibodies, blots were visualized using a chemiluminescence system.

### Confocal microscopy analysis

Confocal microscopy analysis was performed according to standard procedures. In brief, cells were treated with respective agent(s) for 48 hours. Then, the cells were incubated at 37°C for 20 mins in the presence/absence of the BODIPY™ 581/591 C11 (Invitrogen) or FerroOrange (Cayman, #41725). Nuclei were counterstained with 4,6-diamidino-2-phenylindole (DAPI) before mounting. Confocal fluorescence images were captured using an ImageXpress portfolio of high-content image systems (Molecular Devices).

### Mouse model

Animal studies were conducted in accordance with protocols approved by the Institutional Animal Care and Use Committee (IACUC) of China Medical University Hospital. NPG mice were obtained from LASCO Biotechnology Co., Ltd. (LASC, Taiwan) and ASID mice were obtained from the National Laboratory Animal Center (Taiwan). In the PDAC orthotopic xenograft model, 1×10^5^ of CFPAC cells stably expressing luciferase were orthotopically injected into the pancreas of 6-week-old female NPG mice. In the patient-derived xenograft (PDX) model, tumor fragments obtained from a pancreatic cancer patient were subcutaneously implanted into 6-week-old female NPG mice. In the NSCLC orthotopic xenograft models, 5×10^5^ of A549 cells stably expressing luciferase were orthotopically injected into the lungs of 6-week-old male ASID mice. All inhibitors were dissolved in saline. The final concentrations of drugs were imatinib (15 mg/kg) and trametinib (0.3 mg/kg). Treatment was initiated one week after tumor implantation. Mice were treated by oral gavage daily for 30 days, following a schedule of 5 days on treatment and 2 days off. Mouse weight and tumor volume were measured three times every week. In the PDX model, tumor volume was estimated using the following formula: volume (mm^3^) =length (mm)x width (mm)x0.5 width (mm), where length is the longest axis of the tumor. In PDAC and NSCLC orthotopic xenograft models, tumor sizes were monitored using a non-invasive IVIS. Mouse cardiac blood was collected by vegetarians in the Department of Veterinary Medicine and Surgery at National Animal Facility Center after 30 days of treatment, and used to determine the concentrations of alanine aminotransferase, aspartate aminotransferase, and blood urea nitrogen.

### Immunohistochemistry

The mice tissue specimens were collected from the mouse studies. Sample were processed, embedded in paraffin, and sectioned at 3μm. Paraffin sections were deparaffinized and hydrated using the following steps: 5 minutes in xylene three times; 5 minutes in 100% ethanol, 5 minutes in 80% ethanol and 5 minutes in 75% ethanol; and 3 minutes in PBS at room temperature. For p-AKT S473, p-MEK1/2 S217/221, p-ERK1/2 Y204/T202, NCOA4, and HIF-1α staining, 1 mM EDTA buffer (pH 9.0) was used for antigen retrieval. For KI67 staining, Citrate buffer (pH 6.0) was used for antigen retrieval. Endogenous peroxidase was quenched with 3% H_2_O_2_ for 10 minutes, followed by blocking with 3% blocking buffer for 5 minutes. The primary antibodies and their dilutions are as follows: Ki67 (1:250, Invitrogen), p-AKT S473 (1:100, CST, #4060), p-MEK1/2 S217/221 (1:100, CST, #9154), p-ERK1/2 Y204/T202 (1:100, CST, #4370), NCOA4 (1:200, GeneTex, #04250), HIF-1α (1:250, iReal, #IR113-466). The slides were incubated with HRP polymer reagent for 30 minutes. Sections were developed with DAB solution and counterstained with hematoxylin.

### Design and patients in the phase I trial

We conducted a pilot single-cohort clinical trial (ClinicalTrials.gov Identifier: NCT06962254) at China Medical University Hospital. The aim of this study was to assess the safety and preliminary efficacy of a combination regimen (imatinib plus trametinib) in advanced, chemotherapy-refractory PDAC patients with non-G12C *KRAS* mutations. Oral trametinib at a dose of 2 mg and imatinib at a dose of 100 mg were given daily in the morning every 28 days per cycle. Computed tomography for tumor assessment was performed at baseline and every 2 months afterwards, or as clinically indicated. Serum CA19-9 was checked at baseline and every one month after enrollment. Tumor response was assessed according to the Response Evaluation Criteria in Solid Tumors, v1.1 (RECIST v1.1). Treatment was continued until disease progression, occurrence of unacceptable toxicity, death, withdrawal of informed consent, or at the discretion of in-charged physician. Outcome measures included objective response rate (ORR) according to the RECIST v1.1, disease control rate (DCR), progression-free survival (PFS) and overall survival (OS). The DCR was defined as percentage of patients with complete response (CR), partial response (PR) and stable disease (SD). PFS that was calculated from the date of registration to the date of disease progression or death from any cause. OS was calculated from the date of registration until the date of death or the last follow-up. Both PFS and OS were plotted using the Kaplan-Meier method.

### Statistical analysis

All experiments were replated at least three times unless otherwise indicated. Error bar represent standard deviation (SD). Fold changes in Western blot signals were analyzed by a nonparametric Friedman test using GraphPad Prism 9.0 software. P values less than 0.05 were considered statistically significant: *P<0.05, **P<0.01, and ***P<0.0001. Combination index experiments were designed according to the Chou-Talalay method^57^, and results were calculated using CompuSyn software. (http://www.combosyn.com/).

## Data Availability

All data produced in the present study are available upon reasonable request to the authors.

## Notes

### Competing Interest Statement

The authors have declared no competing interest.

### Clinical Trial

NCT06962254

### Author Declarations

Research Ethics Commitee, China Medical University & Hospital

### Funding

- National Science and Technology Council: NSTC 114-2639-B-039-001 -ASP, NSTC 115-2634-F-039-001
- Ministry of Health and Welfare Taiwan: MOHW115-TDU-B-222-114007
- Ministry of Education: The Featured Areas Research Center Program

