## Supplemental data for "Reciprocal Feedback Blockade with Trametinib and Imatinib Overcomes the Limitations of Current KRAS-targeted Therapy"

Supplemental information

This file includes the extended data figure 1~7

Extended data Fig. 1: Multiple tyrosine kinase activation is associated with poor responses to trametinib in *KRAS* mutant cells.

Extended Data Fig. 2: Imatinib modulates MAPK signaling and phosphatase expression in *KRAS* mutant and *KRAS* WT cells

Extended Data Fig. 3: Identification of imatinib as a synergistic partner with trametinib in *KRAS* mutant cells

Extended Data Fig. 4: Combined imatinib and trametinib treatment induces ferritinophagy and lipid peroxidation accumulation

Extended Data Fig. 5: Effects of the combination of imatinib and trametinib on tumor morphology and body weight in *KRAS* mutant xenograft models

Extended Data Fig. 6: Effects of imatinib and trametinib on tumor morphology and body weight in the *KRAS* mutant PDX model

Extended Data Fig. 7: Clinical activity of combined imatinib and trametinib in participants with *KRAS* mutant tumors

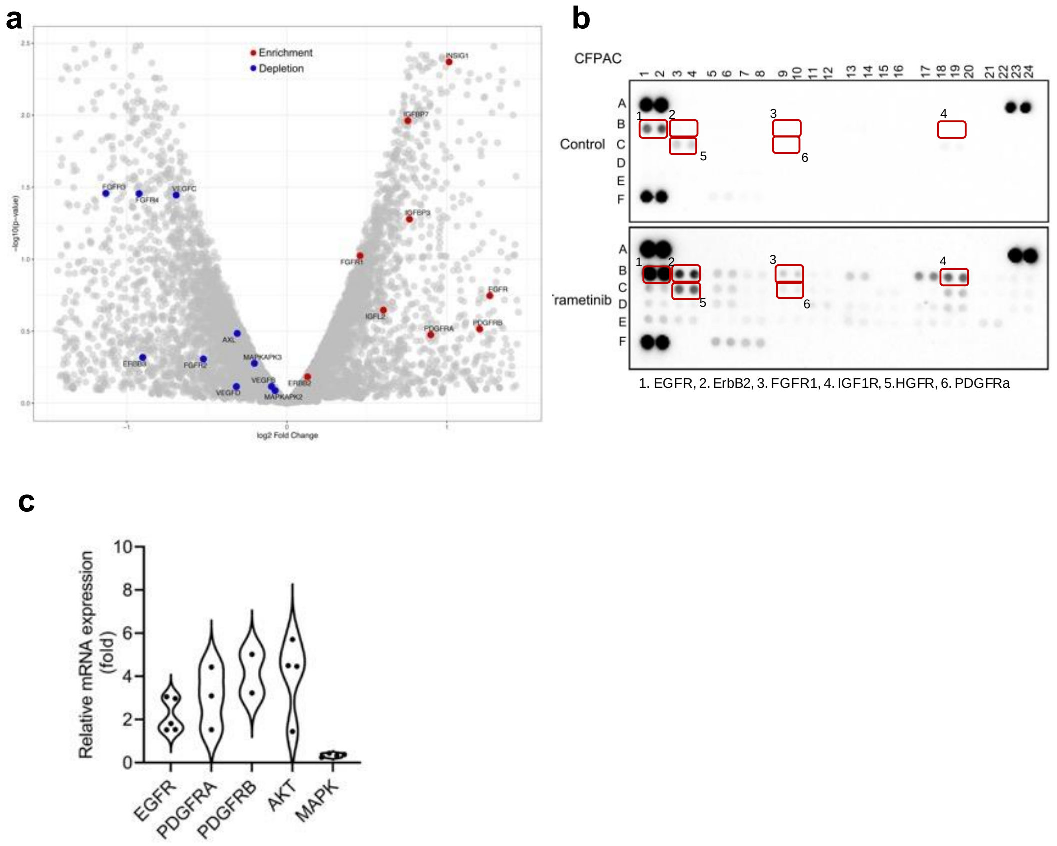


**Extended Data Fig. 1: Multiple tyrosine kinase activation is associated with poor responses to trametinib in *KRAS* mutant cells**

**a,** Scatter plot showing $\log_2$ fold changes in trametinib-treated cells compared to the untreated control, and RTK-and angiogenesis-related genes are shown in red (upregulated) and blue (downregulated). **b,** RTK antibody array analysis was performed using cell lysates of CFPAC cells treated/untreated with trametinib (30 nM) for 48 hours. Signals were detected for phosphorylated EGFR, ErbB2, FGFR1, insulin-like growth factor (IGF-R), hepatocyte growth factor receptor (HGFR/MET) and PDGFRα. **c,** The relative mRNA expression levels of *EGFR*, *PDGFRA*, *PDGFRB*, *AKT*, and *MAPK*-related genes in A549 cells treated with trametinib (30 nM) for 24 hours compared to untreated cells were determined by qPCR analysis.

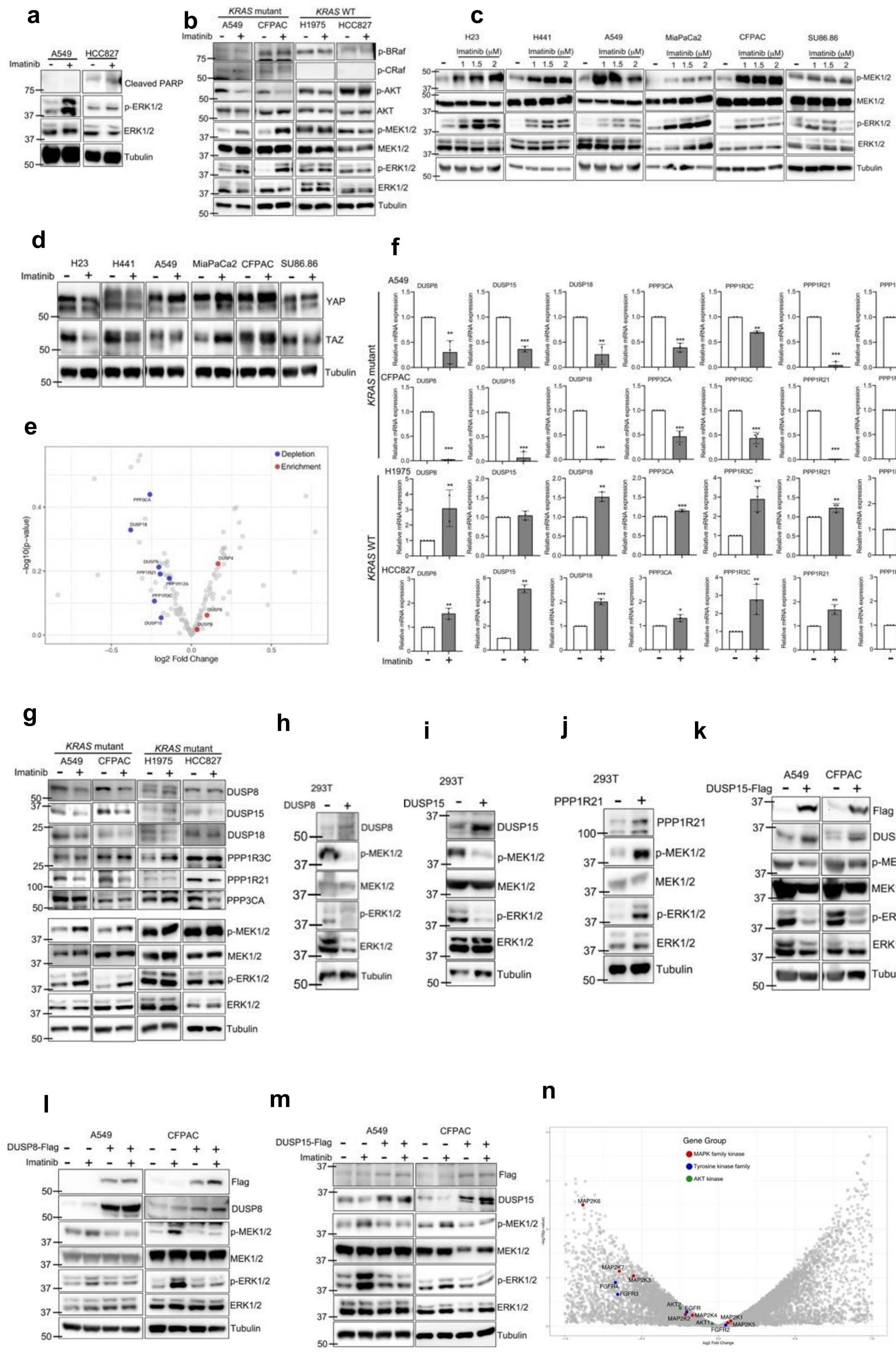

a
A549
HCC827
Imatinib
Cleaved PARP
p-ERK1/2
ERK1/2
Tubulin
b
KRAS mutant
KRAS WT
A549
CFPAC
H1975
HCC827
p-BRaf
p-CRaf
p-AKT
AKT
p-MEK1/2
MEK1/2
p-ERK1/2
ERK1/2
Tubulin
c
H23
H441
A549
MiaPaCa2
CFPAC
SU86.86
Imatinib (μM)
p-MEK1/2
MEK1/2
p-ERK1/2
ERK1/2
Tubulin
d
H23
H441
A549
MiaPaCa2
CFPAC
SU86.86
Imatinib
YAP
TAZ
Tubulin
e
Depletion
Enrichment
−log10(p−value)
log2 Fold Change
PPP3CA
DUSP18
DUSP8
PPP1R21
PPP1R12A
PPP1R3C
DUSP15
DUSP4
DUSP6
DUSP9
f
KRAS mutant
KRAS WT
A549
CFPAC
H1975
HCC827
DUSP8
DUSP15
DUSP18
PPP3CA
PPP1R3C
PPP1R21
PPP1R12A
Relative mRNA expression
Imatinib
g
KRAS mutant
A549
CFPAC
H1975
HCC827
Imatinib
DUSP8
DUSP15
DUSP18
PPP1R3C
PPP1R21
PPP3CA
p-MEK1/2
MEK1/2
p-ERK1/2
ERK1/2
Tubulin
h
293T
DUSP8
p-MEK1/2
MEK1/2
p-ERK1/2
ERK1/2
Tubulin
i
293T
DUSP15
p-MEK1/2
MEK1/2
p-ERK1/2
ERK1/2
Tubulin
j
293T
PPP1R21
p-MEK1/2
MEK1/2
p-ERK1/2
ERK1/2
Tubulin
k
A549
CFPAC
DUSP15-Flag
Flag
DUSP15
p-MEK1/2
MEK1/2
p-ERK1/2
ERK1/2
Tubulin
l
A549
CFPAC
DUSP8-Flag
Imatinib
Flag
DUSP8
p-MEK1/2
MEK1/2
p-ERK1/2
ERK1/2
Tubulin
m
A549
CFPAC
DUSP15-Flag
Imatinib
Flag
DUSP15
p-MEK1/2
MEK1/2
p-ERK1/2
ERK1/2
Tubulin
n
Gene Group
MAPK family kinase
Tyrosine kinase family
AKT kinase
MAP2K6
MAP2K7
MAP2K3
FGFR4
FGFR3
AKT3
EGFR
MAP2K2
MAP2K4
AKT1
MAP2K1
MAP2K5
FGFR2
log2 Fold Change

**Extended Data Fig. 2: Imatinib modulates MAPK signaling and phosphatase expression in *KRAS* mutant and *KRAS* WT cells**

**a,** Western blot analysis of cleaved PARP, p-ERK1/2 (Y204/T202) and ERK1/2 in A549 and HCC827 cells treated with 10 µM of imatinib for 24 hours. **b,** A549, CFPAC, H1975 and HCC827 cells were treated/untreated with imatinib (1 µM) for 24 hours, and S445-phosphorylated BRaf (p-BRaf), S338-phosphorylated CRaf (p-CRaf), p-AKT (S473), AKT, p-MEK1/2 (S217/221), MEK1/2, p-ERK1/2 (Y204/T202), and ERK1/2 were analyzed by western blot. **c,** *KRAS* mutant NSCLC H23 (*KRAS* $^{G12C}$), H441 (*KRAS* $^{G12V}$), A549 (*KRAS* $^{G12S}$) and PDAC MiaPaCa-2 (*KRAS* $^{G12C}$), CFPAC (*KRAS* $^{G12V}$), SU.86.86 (*KRAS* $^{G12D}$) cells were treated with imatinib at the indicated concentrations for 24 h, followed by western blot analysis of p-MEK1/2 (S217/221), MEK1/2, p-ERK1/2 (Y204/T202), and total ERK1/2. **d,** H23, H441, A549, MiaPaCa-2, CFPAC, and SU.86.86 cells were treated with imatinib (1 µM) for 24 h, and YAP, and TAZ expression were analyzed by western blot. **e,** Scatter plot showing $\log_2$ fold changes in gene expression in 1 µM imatinib-treated versus control samples. Phosphatase genes are shown in red (upregulated) and blue (downregulated). **f,** qRT-PCR analysis of *DUSP8*, *DUSP15*, *DUSP18*, *PPP3CA*, *PPP1R3C*, *PPP1R21* and *PPP1R12A* in A549, CFPAC, H1975 and HCC827 cells after 6 hours of 1µM imatinib treatment. Data represent mean ± SD from at least three independent experiments. *p < 0.05, **p < 0.01,***<0.001. **g,** Western blot analysis of DUSP8, DUSP15, of DUSP18, PPP1R3C, PPP1R21 and PPP3CA, p-MEK1/2(S217/221), MEK1/2, p-ERK1/2 (Y204/T202), and ERK1/2 in A549, CFPAC, H1975 and HCC827 cells treated/untreated with 1µM imatinib. **h-j,** Western blot analysis of DUSP8, DUSP15, PPP1R21 p-MEK1/2 (S217/221), MEK1/2, p-ERK1/2 (Y204/T202) and ERK1/2 in 293T cells overexpressing DUSP8 (**h**), DUSP15 (**i**), or PPP1R21 (**j**) via viral transduction. **k,** Western blot analysis of Flag-tag, DUSP15, p-MEK1/2 (S217/221), MEK1/2, p-ERK1/2(Y204/T202) and ERK1/2 in A549 and CFPAC cells overexpressing DUSP15. **l,m,** Western blot analysis of Flag-tag, DUSP8 (**l**), DUSP15 (**m**), p-MEK1/2 (S217/221), MEK1/2, p-ERK1/2 (Y204/T202) and ERK1/2 in A549 and CFPAC cells overexpressing DUSP8 or DUSP15 following 6 hours of 1µM imatinib treatment. **n,** Scatter plot showing $\log_2$ fold changes in gene expression following the combination of imatinib and trametinib treatment and MAPK-family, tyrosine kinases, and AKT genes are shown in red, blue and green, respectively.

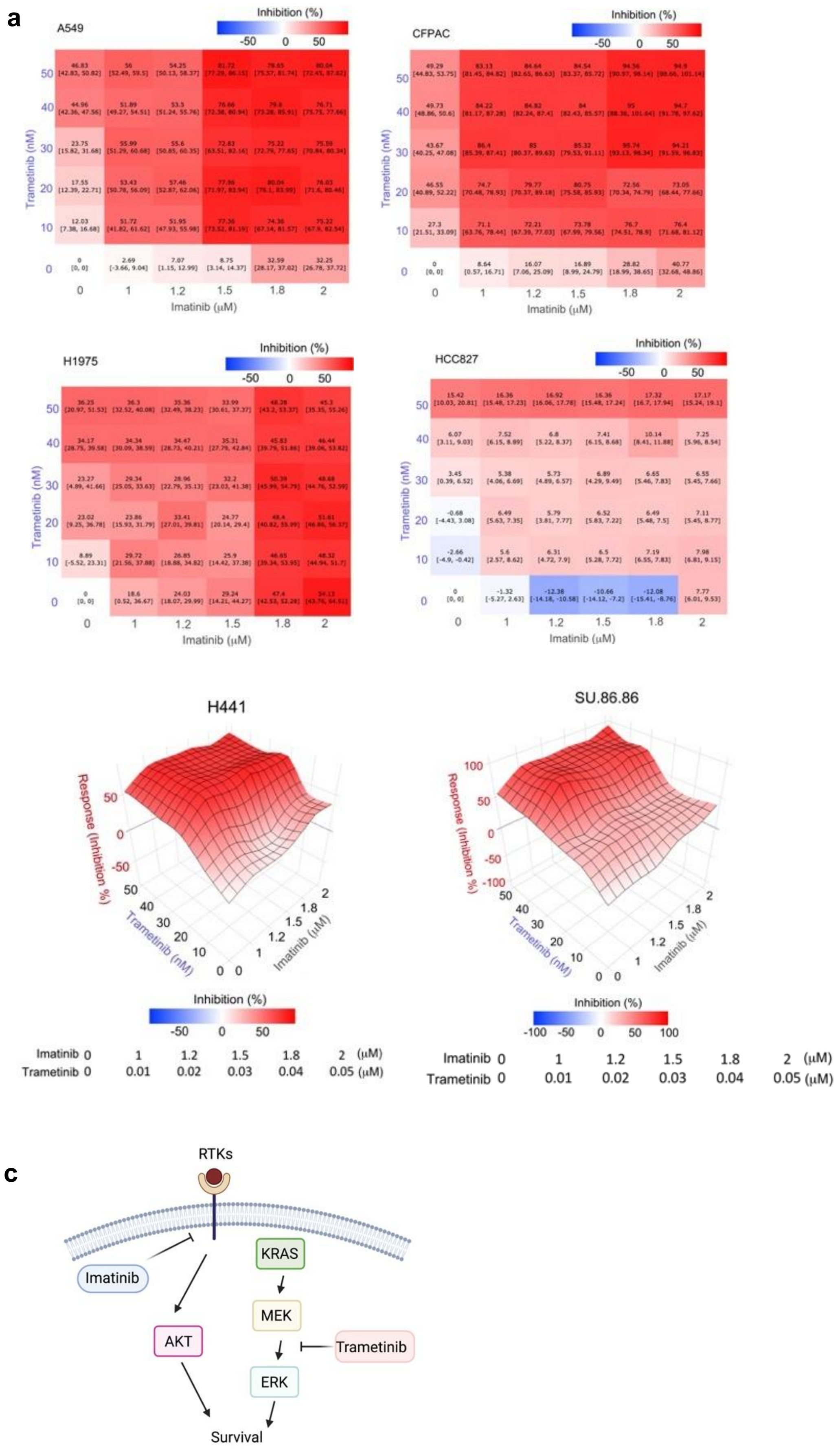
a
A549
CFPAC
H1975
HCC827
Inhibition (%)
-50
0
50
Trametinib (nM)
Imatinib (μM)
b
H441
SU.86.86
Response (Inhibition %)
Trametinib (nM)
Imatinib (μM)
Inhibition (%)
-100
-50
0
50
100
Imatinib 0 1 1.2 1.5 1.8 2 (μM)
Trametinib 0 0.01 0.02 0.03 0.04 0.05 (μM)
c
RTKs
Imatinib
KRAS
MEK
AKT
ERK
Trametinib
Survival

**Extended Data Fig. 3: Identification of imatinib as a synergistic partner with trametinib in *KRAS* mutant cells**

**a,** Heatmap depicting the effect of trametinib and imatinib on *KRAS* mutant cells, A549, CFPAC, and *KRAS* WT H1975 and HCC827 cells. Cells were treated with the indicated concentrations for 6 days, and cell viability was assessed by colony formation assay. **b,** Synergistic anti-cancer effects of trametinib and imatinib in *KRAS* mutant cells, H441 and SU.86.86 cells under the same conditions. **c,** Schematic model illustrating the mechanism underlying the synergistic effect of combined imatinib and trametinib treatment in *KRAS* mutant cells.

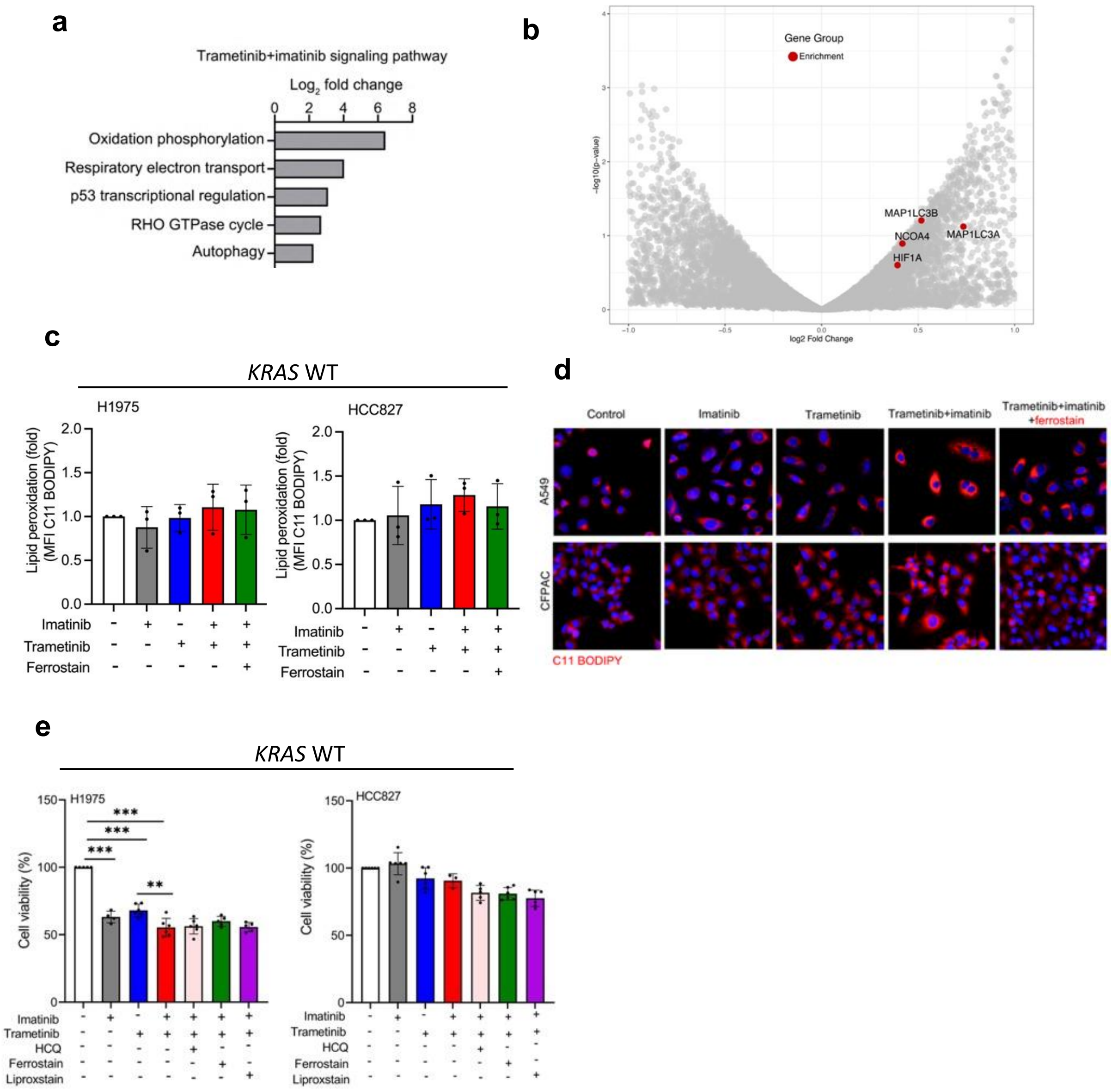


**Extended Data Fig. 4: Combined imatinib and trametinib treatment induces ferritinophagy and lipid peroxidation accumulation**

**a,** Bar graph of pathway analysis using bioplanet dataset analysis in RNA-sequencing data of cells treated with the imatinib-trametinib combination compared with the control for 72 h. **b,** Scatter plot showing $log_2$ fold changes of genes in cells treated with the combination of imatinib and trametinib compared to untreated cells. The ferritionophagy-related genes are shown in red. **c,** The levels of lipid peroxidation in H1975 and HCC827 cells treated with 1µM imatinib and/or 30nM trametinib in the presence or absence of 1 µM ferrostatin for 72 h. Data represent mean ± SD from at least three independent experiments. **d,** A549 and CFPAC cells were treated with 1 µM imatinib and/or 30 nM trametinib in the presence/absence of 1 µM ferrostatin for 72 h, and lipid peroxidation detected by BODIPY C11 analyzed by confocal immunofluorescence staining (red). **e,** Colony formation assay of H1975 and HCC827 cells treated with 1 µM imatinib and/or 30 nM trametinib in the presence/absence of 10 µM hydroxychloroquine (HCQ), 1 µM ferrostatin, or 2 µM liproxstatin for 6 days. Data represent mean ± SD from at least three independent experiments. $^{**}p < 0.01$.

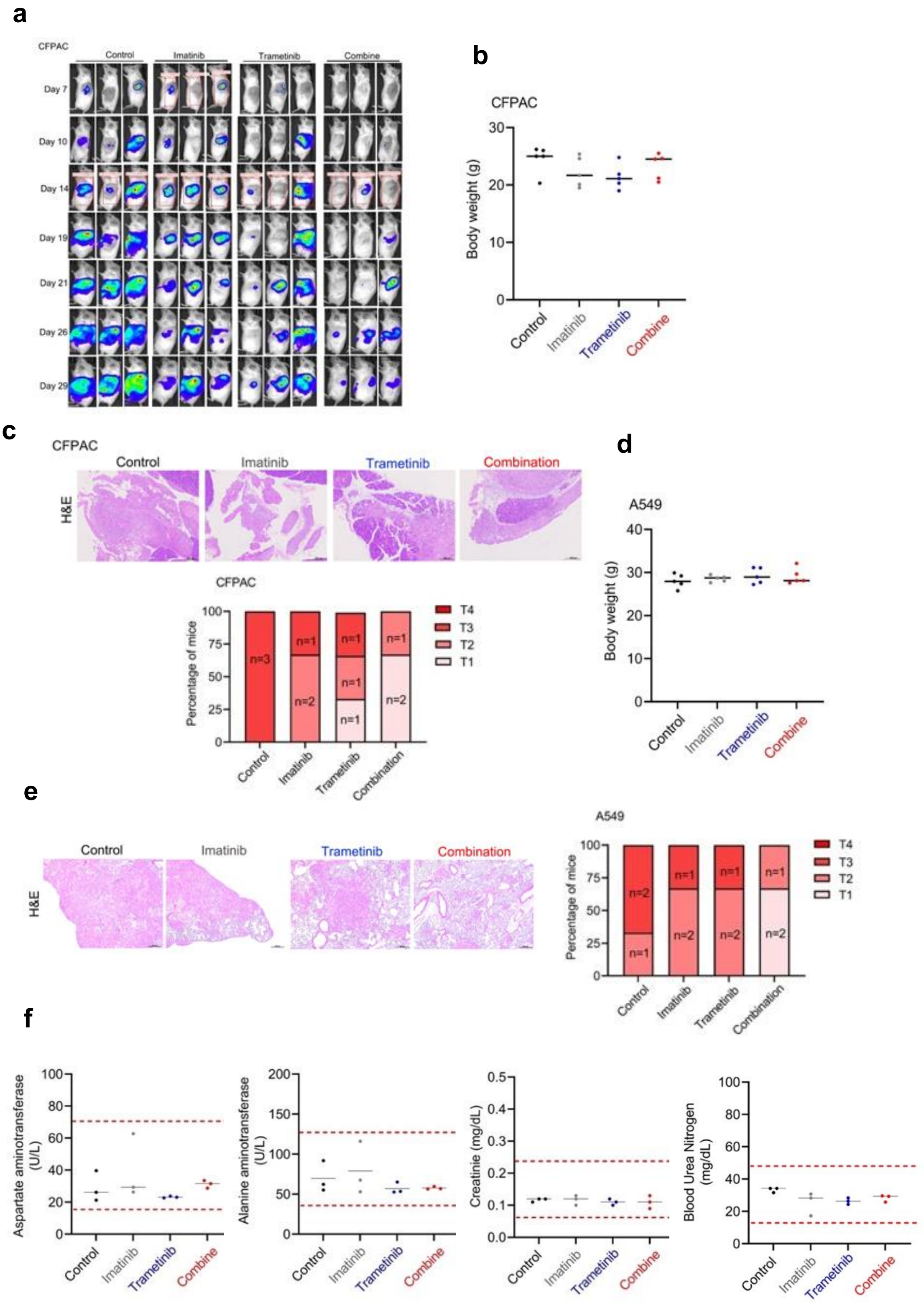


**Extended Data Fig. 5: Effects of the combination of imatinib and trametinib on tumor morphology and body weight in *KRAS* mutant xenograft models**

**a,** Representative luminescence imaging of the mice used in Fig. 4a-c over time following oral treatment with vehicle, imatinib, trametinib, or the combination. **b,**

Body weight of the mice on day 30 used in Fig. 4a. Data represent mean ± SEM; n = 5 mice/group. **c,** Representative H&E staining (4×) and T classification quantification of the tumors from the mice on day 30 used in Fig 4a. **d,** Body weight of the mice on day 30 used in Fig 4f, g. Data represent mean ± SEM; n = 5 mice/group. **e,** Representative H&E staining (4×) and T classification quantification of the tumors from the mice on day 30 used in Fig. 4f. **f,** Hepatotoxicity, AST and ALT and renal toxicity, creatinine and BUN in ASID mice bearing *KRAS* G12S-mutant xenografts treated with imatinib, trametinib, or the combination for 30 days. Data presented mean ± SEM; n = 3 mice/group.

**a**

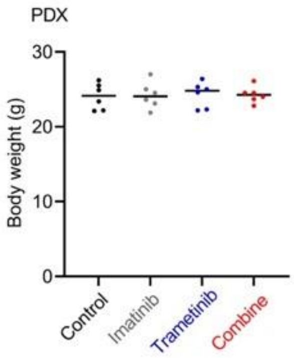


**b**

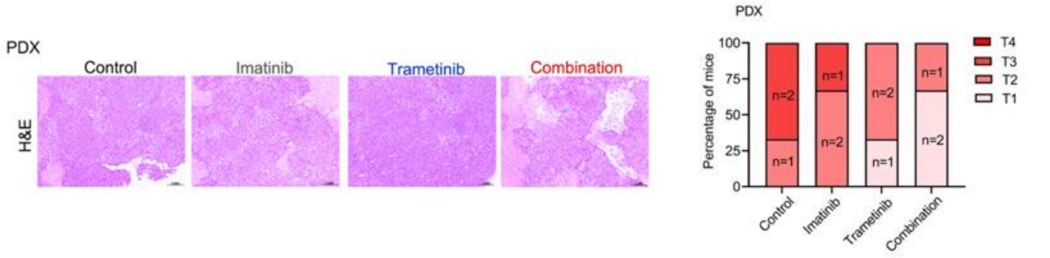


**Extended Data Fig. 6: Effects of imatinib and trametinib on tumor morphology and body weight in the *KRAS* mutant PDX model**

**a,** Body weight of the mice on day 30 used in Fig 5c. Data represent mean ± SEM; n = 6 mice/group. **b,** Representative H&E staining (4×) and quantification percentage of T classification of the tumors from the mice on day 30 used in Fig. 5c.

a

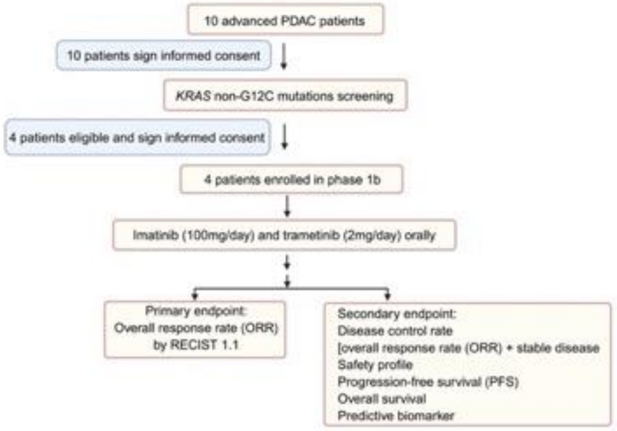


b

| Participants | #1 | #2 | #3 | #4 |
|---|---|---|---|---|
| Gender | Female | Female | Male | Female |
| Age | 73 | 64 | 63 | 75 |
| Pathology type | Adenocarcinoma | Adenocarcinoma | Adenocarcinoma | Adenocarcinoma |
| KRAS mutation | *KRAS* G12R | *KRAS* G12V | *KRAS* G12D | *KRAS* G12R |
| Stage | Stage IV | Stage IV | Stage IV | Stage IV |
| Stage at diagnosis | cT3N1M1 | cT4N1M1 | pT2N1M0 | cT2N1M0 |
| Prior treatment line | 1 | 1 | 2 | 2 |
| State at enrollment | Peritoneal carcinomatosis | Liver and lymph node metastasis | lymph node and lung metastasis | Liver metastasis |

c

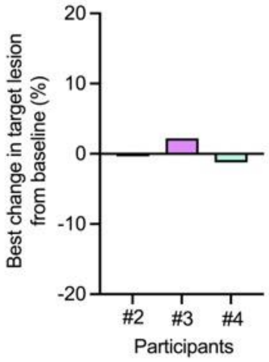


**Extended Data Fig. 7: Clinical activity of combined imatinib and trametinib in participants with *KRAS* mutant tumors**

**a,** Schema evaluating imatinib plus trametinib in participants with *KRAS*-mutant tumors. **b,** Baseline clinical characteristics of the four participants (#1–#4). **c,** Best tumor response.
